# Thiamine pyrophosphokinase-1 deficiency in neurons drives Alzheimer’s multiple pathophysiological alterations

**DOI:** 10.1101/2024.09.20.24314010

**Authors:** Shaoming Sang, Xiangteng Zhao, Ting Qian, Yingfeng Xia, Xiaoli Pan, Qianhua Zhao, Fang Cai, Yeting Zeng, Wenwen Cai, Boru Jin, Hongyan Qiu, Yangqi Xu, Qiang Huang, Yun Zhang, Shajin Huang, Donglang Jiang, Yun Wu, Haiyang Tong, Qing Zhang, Changpeng Wang, Xiaoqin Cheng, Kai Zhong, Yihui Guan, Michael X. Zhu, Xiang Yu, Peng Yuan, Weihong Song, Chunjiu Zhong, Benfotiamine Phase 2 Clinical Trial Investigators

## Abstract

**Background:** The mechanism driving multiple pathophysiological alterations in Alzheimer’s disease (AD) remains unclear. Thiamine deficiency, a well-known feature of AD, may contribute to these alterations.

**Methods:** The expressions of four known genes associated with thiamine metabolism were studied in brain samples from patients with AD and other neurodegenerative disorders. The results were further demonstrated in AD and diabetic mouse and cellular models. The phenotypes of mice with conditional *Thiamine pyrophosphokinase-1* (*Tpk*) knockout in brain excitatory neurons were investigated. The therapeutic effects of thiamine diphosphate supplement and *Tpk* delivery on cellular and mouse models were explored. Phase 2 clinical trial of benfotiamine, a thiamine derivative, plus donepezil was performed.

**Results:** Only TPK expression was inhibited in brain samples of AD patients, while none of thiamine-associated genes were significantly changed in other neurodegenerative disorders. TPK inhibition in the brains and neurons was verified in AD and diabetic mouse and cellular models. Mice with *Tpk* deletion in neurons exhibited all major pathophysiological alterations of AD, including amyloid deposition, Tau hyperphosphorylation, and brain atrophy. TPK expression restoration and thiamine diphosphate supplement ameliorated the pathophysiological and behavioral phenotypes in mouse and cell models with *Tpk* insufficiency. Benfotiamine delayed cognitive decline in mild-to-moderate AD patients with a dose-effect relationship, particularly with a significant attenuation of the deterioration in moderate AD patients by post hoc analysis.

**Conclusions:** TPK deficiency and hence thiamine diphosphate reduction in neurons are a decisive factor driving multiple pathophysiologic alterations of AD, unveiling a new direction for the disease mechanism and treatment.

## Introduction

Alzheimer’s disease (AD) is a complex neurodegenerative disorder, which involves two biologic stages, numerous risk factors, and multiple pathophysiological features. Two biological stages refer to the preclinical stage and the symptomatic stage. The former lasts for 10 to 20 years and is mainly characterized by amyloid-β (Aβ) deposition. The latter is characterized by multiple pathophysiological features and progresses relatively rapidly. In addition to Aβ deposition, multiple pathophysiological features of AD include glial activation and neuroinflammation, abnormal glucose metabolism, Tau hyperphosphorylation aggregating to the tangles, progressive loss of synapses and neurons leading to brain atrophy, and so forth. The risk factors of AD can be grouped into aging, genetic factors, chronic diseases or pathological conditions, unhealthy lifestyles, and educational status.

Brain Aβ deposition is an indispensable feature and the earliest biomarker with detectable change of AD ^1-3^. Further, the mutations of three genes, *APP*, *PSEN1*, and *PSEN2*, cause dominantly inherited cases by enhancing the production and aggregation of Aβ ^4-8^. These facts strongly indicate that brain Aβ deposition contributes to the pathogenesis of AD ^9, 10^. However, the mechanism by which Aβ deposition induces other multiple pathophysiological alterations, especially neurodegenerative pathology, still needs to be clarified.

Brain glucose hypometabolism is an invariant pathophysiological feature and indicator of neurodegeneration in AD, which closely correlates with the degree of cognitive impairment and the progression of the disease ^11, 12^. The previous studies have demonstrated that the blood and brain samples of AD patients exhibit significant reduction in the activities of three key enzymes involved in intracellular glucose metabolism: pyruvate dehydrogenase and α-ketoglutarate dehydrogenase in the Krebs cycle and transketolase in the non-oxidative branch of pentose phosphate pathway ^13-16^. These three enzymes have a common coenzyme: thiamine diphosphate (TDP). Our previous studies have demonstrated that TDP reduction is significant and closely correlates with cerebral glucose hypometabolism in AD ^17, 18^. Further, thiamine deficiency by diet deprivation significantly exacerbates brain Aβ deposition in an AD mouse model ^19^. Therefore, thiamine deficiency may contribute to the occurrence and development of many of the multiple pathophysiological alterations in AD.

There are four known genes associated with thiamine metabolism, including *Thiamine Pyrophosphokinase-1* (*TPK*), *Solute Carrier Family 19 Member 2* (*SLC19A2*), *SLC19A3*, and *SLC25A19*. *TPK* is responsible for converting thiamine into functional TDP in cytoplasm, while the other three genes contribute to the transportation of thiamine and TDP on cell or mitochondria membrane. In the current studies, we first investigated the expressions of these four genes in brain samples of patients with AD and other neurodegenerative diseases. The effects of Aβ plaques and oligomers as well as diabetes, a well-known risk factor of sporadic AD ^20, 21^, on TPK expression were further demonstrated. Then, the phenotypes in adult mice with conditional *Tpk* knockout in brain excitatory neurons and corresponding rescue efficacies were analyzed. Finally, a phase 2 clinical trial of benfotiamine, a thiamine derivative that significantly elevates TDP levels in human blood and erythrocytes ^22-24^, was performed.

## Methods

### Human study

The mRNA levels of the four known genes in brain samples of patients with AD and other neurodegenerative disorders in multiple independent public datasets were analyzed. Brain TPK protein level of AD patients and control subjects were further examined using Western blot analysis.

### Studies on mouse and cell models

The effects of Aβ plaques and oligomers and high glucose on the expression of TPK were explored in mouse and cell models. A model of conditional *Tpk* knockout in brain excitatory neurons of adult mice (*Tpk*-cKO mice) was established. Biochemical and morphological studies were performed using Western blotting, ELISA, high performance liquid chromatography, untargeted metabolomics, immunofluorescent and Nissl staining, structural T2-weighted magnetic resonance imaging, silver staining, and scanning and transmission electron microscopy of mouse brains. Positron emission tomography / computer tomography with ^18^F-fluorodeoxyglucose (FDG-PET / CT) and behavior tests were applied. Adeno-associated virus-mediated delivery of *Tpk* gene and thiamine or TDP supplement treatment was employed to determine their rescuing effects on pathophysiological features and behavior abnormality (Details in Supplementary Methods and Materials).

### A multicenter, randomized, phase 2 clinical trial

A randomized, double-blind, placebo-controlled, 52-week trial (chiCTR18000143167) of high dose (600 mg daily) and low dose (300 mg daily) benfotiamine plus donepezil (5 mg daily) treatment was conducted in patients with mild-to-moderate AD, defined based on a score of 11 to 24 on the Mini-Mental State Examination (MMSE, score range: 0 to 30, with higher scores indicating better cognitive abilities) and on the diagnostic guideline for AD (the National Institute on Aging-Alzheimer’s Association workgroups) ^25^. All patients must have taken donepezil (5 mg daily) continuously for more than six months before registration. The primary endpoint was the changes at week 52 from the baseline in the scores of AD Assessment Scale-Cognitive subscale 11 (ADAS-cog, score range: 0 to 70, with higher scores indicating more severe cognitive impairment). The key secondary endpoint outcomes included the changes of MMSE and ADAS-activities of daily living (ADAS-ADL) scores at week 52 from the baseline. The missing data of efficacy indicators, including the scores of ADAS-cog, MMSE, and ADAS-ADL, were imputed using the last observation carried forward (LOCF). The statistical analysis used the mixed model for repeated measures (MMRM) of the SAS software (v.9.4). More details presented in Supplementary Methods and Materials.

## Results

### Specific down-regulation of TPK expression in brains of AD patients

The mRNA levels of the four known genes were first analyzed in multiple independent public datasets of patients with AD and other neurodegenerative diseases. In the Illumina HiSeq 2500 RNA-seq dataset (GSE95587) that measured the fusiform gyrus samples ^26^, *TPK* mRNA levels in AD patients were significantly reduced as compared with that in control subjects with normal cognition (NC), while mRNA levels of the other three genes (*SLC19A2*, *SLC19A3*, and *SLC25A19*) showed no significant change (Fig. 1A; Table S1). *TPK* mRNA levels were found to be negatively correlated with Braak staging scores in AD patients, while the correlation tended to be positive in NC subjects (R = 0.31, P = 0.079, Fig. 1B). Further, for subjects with Braak stage □ pathology, NC subjects exhibit significantly higher *TPK* mRNA levels than AD patients (Fig. 1C). Those results indicated that higher *TPK* mRNA level may maintain cognitive function in the asymptomatic stage even with significant neuropathologic burdens. The results imply that the insufficiency of TPK expression is closely associated with symptom onset of AD.

**Fig 1.**
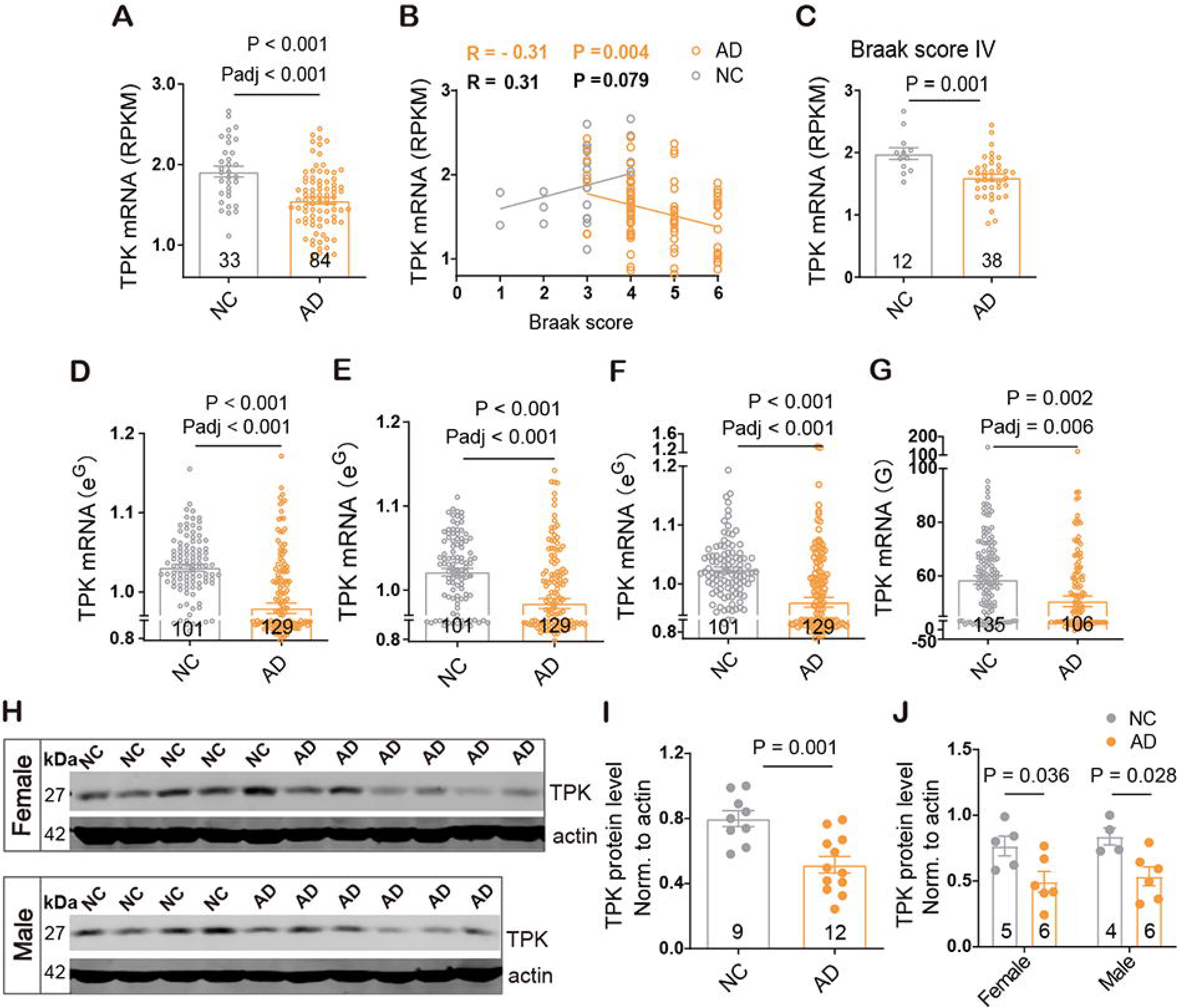
Specific down-regulation of TPK expression in brains of AD patients. Panel A shows *TPK* mRNA levels in fusiform gyrus cortices of AD patients from the GSE95587 dataset were significantly lower than that in control subjects with normal cognition (NC). Panel B shows correlation between brain *TPK* mRNA levels and scores of Braak staging: while the correlation was negative for AD patients (n = 84), it tended to be positive for control subjects (n = 33). Panel C shows for individuals with Braak stage IV pathology, NC subjects showed significantly higher levels of *TPK* mRNA than AD patients. Panels D-G show TPK mRNA levels were significantly reduced in AD patients compared to NC subjects in the GSE15222 dataset measuring frontal cortex samples (D), the GSE44768 dataset measuring cerebellum samples (E), the GSE44770 dataset measuring dorsolateral prefrontal cortex samples (F), and the GSE44771 dataset measuring visual cortex samples (G). Panels H and I show TPK protein levels in frontal cortices of AD patients were significantly lower than that in NC subjects (P = 0.001). Panel J shows TPK protein levels in frontal cortices of male and female AD patients were significantly lower than that in respective NC subjects (Male: P = 0.036; Female: P = 0.028; P = 0.999 for male *vs*. female AD cases; P = 0.990 for male *vs*. female NC subjects).Statistical analyses were performed using unpaired two-tailed Student’s t-tests, one-way analysis of variance (ANOVA) or Pearson correlation analysis. Summary data represent means ± SEM. P values and adjusted P values (Padj), determined using Benjamini-Hochberg method, are indicated above the bars or > 0.05 if not labeled. The number of human subjects per group is shown in the graph.

The reduction of *TPK* mRNA levels in AD patients was further analyzed in other datasets, including GSE15222 ^27^, GSE44768, GSE44770, and GSE44771 ^28^, while the mRNA levels of other three genes exhibited either no significant change or a slight enhancement (Fig. 1D-G; Table S1). There were no significant changes in the mRNA levels of *TPK*, *SLC25A19*, *SLC19A2*, and *SLC19A3* in brain samples of patients with other neurodegenerative diseases, including frontotemporal dementia (FTLD), Parkinson’s disease (PD), Huntington’s disease (HD), and amyotrophic lateral sclerosis (ALS, Fig. S1A-G, Table S1) ^29-32^. These results demonstrate that the reduced *TPK* expression is specific to AD.

The TPK protein level in cortical samples of AD patients and control subjects of NC (Demographic data in Table S1) was further investigated. The results showed significantly lower TPK protein level in AD patients than NC subjects (Fig. 1H, I). Male and female patients had similar reductions of TPK protein levels (Fig. 1J).

### Aβ deposition and high glucose obviously down-regulate TPK expression in neurons

The effects of Aβ deposition on TPK expression was firstly studied in brain samples of 2, 8, and 16-month-old APP / PS1 transgenic mice (Fig. S2A), which the mRNA and protein levels of TPK were not significantly changed as compared with those in control littermates (Fig. S2B, C). Immunofluorescent staining of TPK showed a significant reduction in the level of TPK in neurons close to the plaques (Fig. 2A).

**Fig 2.**
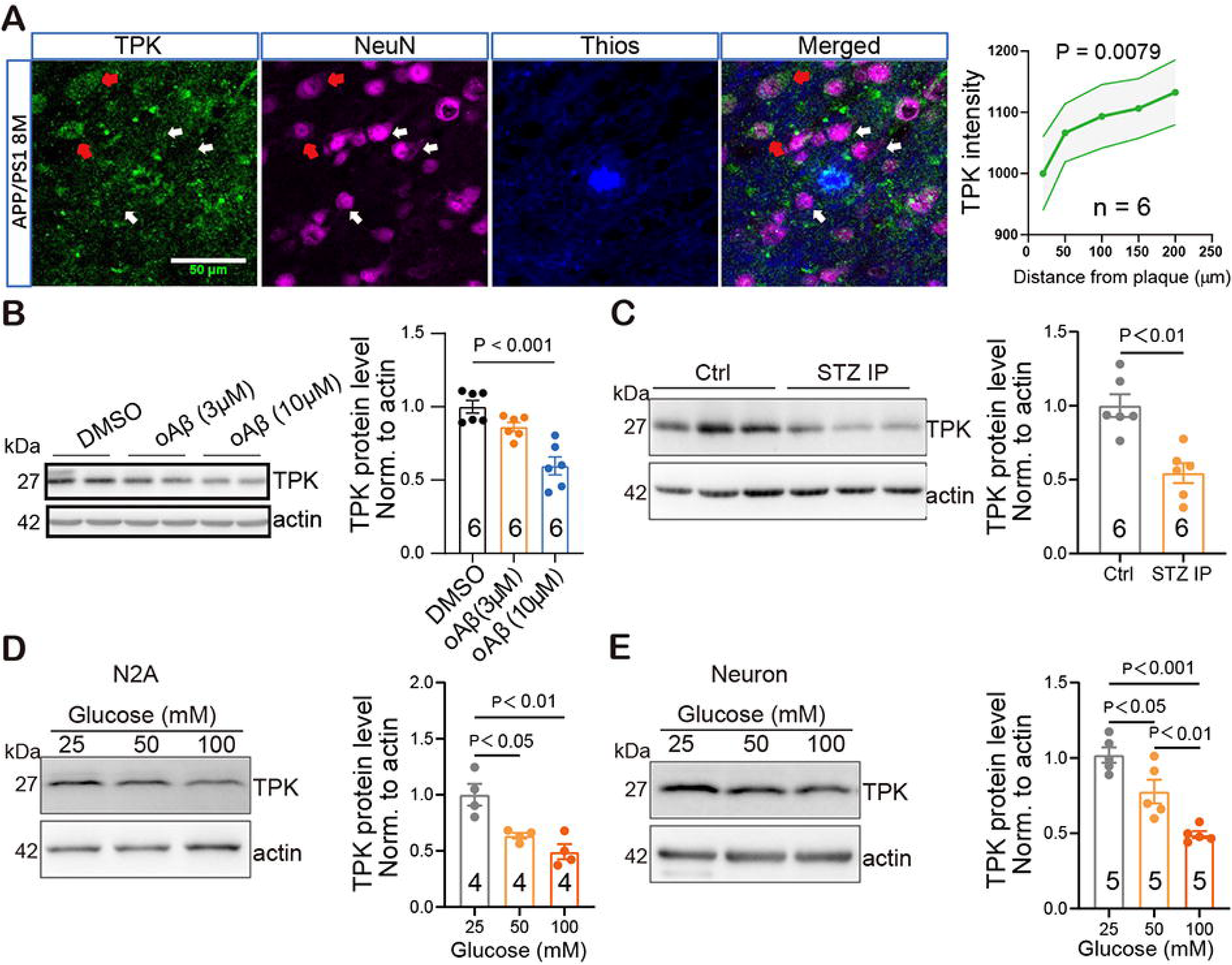
Amyloid-β deposition and diabetes down-regulate TPK expression in neurons. Panel A shows representative images and quantifications of the TPK intensities in neurons near and far away from the plaques in cortices of APP/PS1 transgenic mice, Red arrows indicate high TPK expression in neurons away from the plagues, white arrows indicate low TPK expression in neurons near the plagues. Scale bars, 50 µm. Panel B shows images of Western blots and quantification of TPK protein. Oligomer Aβ dose-dependently decreased the level of TPK protein in primary neurons. Panel C shows images of Western blots and quantification of TPK protein. The level of TPK protein was significantly reduced in cortices of mice 3 days after intraperitoneal STZ injection (STZ IP, 200 mg / kg) as compared with that in control mice. Panels D and E show that high glucose significantly decreased the level of TPK protein in N2a cells (D) and primary neurons (E). Summary data represent means ± SEM. P values are indicated above the bars or > 0.05 if not labeled.

Oligomeric Aβ is well recognized as the most important component of neurotoxicity for Aβ pathology. The effects of oligomeric Aβ on TPK expression in neurons *in vitro* were performed. The results showed that oligomeric Aβ dose-dependently inhibited TPK expression in primary neurons and Neuro-2a (N2a) cells (Fig. 2B; Fig. S3A). Importantly, the elevations of neuron death and neurodegeneration markers, Cleaved-PARP1 and Cleaved-Caspase3, and p-NF-H, induced by oligomeric Aβ were blocked by TPK re-expression in the above cells (Fig. S3B, C).

Diabetes is a well-known independent risk factor of sporadic AD ^20, 21^. To investigate whether diabetes induces TPK inhibition in neurons or not, diabetic mouse and cellular models were exploited. In the conventional mouse model of drug-induced diabetes, blood glucose level was significantly increased three days after intraperitoneal streptozotocin (STZ) injection (Fig. S4A). Among the four known genes associated with thiamine metabolism, only *Tpk* expression was significantly inhibited in the cortices, but not livers at both the protein and mRNA levels (Fig. 2C; Fig. S4B-F).

In cultured primary neurons and N2a cells, the treatment of high glucose, mimicking hyperglycemia, significantly decreased TPK expression in a concentration-dependent manner, which was not seen in the non-neuronal HEK293 cells (Fig. 2D, E; Fig. S4G, H). Together, these results indicate that high glucose, the prominent feature of diabetes, inhibits TPK expression in a tissue- and cell-type selective manner, with the brain and neurons being specifically vulnerable to high glucose.

### Establishment of a model with neuronal *Tpk* knockout in adult mice

To examine the pathophysiological effects of neuronal TPK deficiency, a mouse model was established, in which the *Tpk* gene was conditionally knocked out from glutamatergic neurons of cerebral cortex and hippocampus of adult mice by crossing the tamoxifen-inducible *CaMKII-Cre^ERT2/+^*mice ^33^ with the *Tpk*^fl/fl^ mice (*Tpk*-cKO mice, Fig.3A; Fig. S5A-F). At 12 weeks old, both the *Tpk-*cKO mice and control littermates (*Tpk^fl/fl^*) were intraperitoneally injected with tamoxifen (50 mg / kg / day) for 5 consecutive days. The animals were sacrificed in the 4^th^, 6^th^, 8^th^, or 10^th^ weeks after tamoxifen treatment for biochemical and pathological analysis (Fig. 3A). When measured in the 10^th^ week after tamoxifen treatment, the levels of TPK protein and TDP were significantly reduced in brain samples of the *Tpk*-cKO mice compared to their control littermates (Fig. 3B, C; Fig. S6A, B). Blood TDP levels were not significantly affected in the *Tpk*-cKO mice (Fig. S6C).

**Fig 3.**
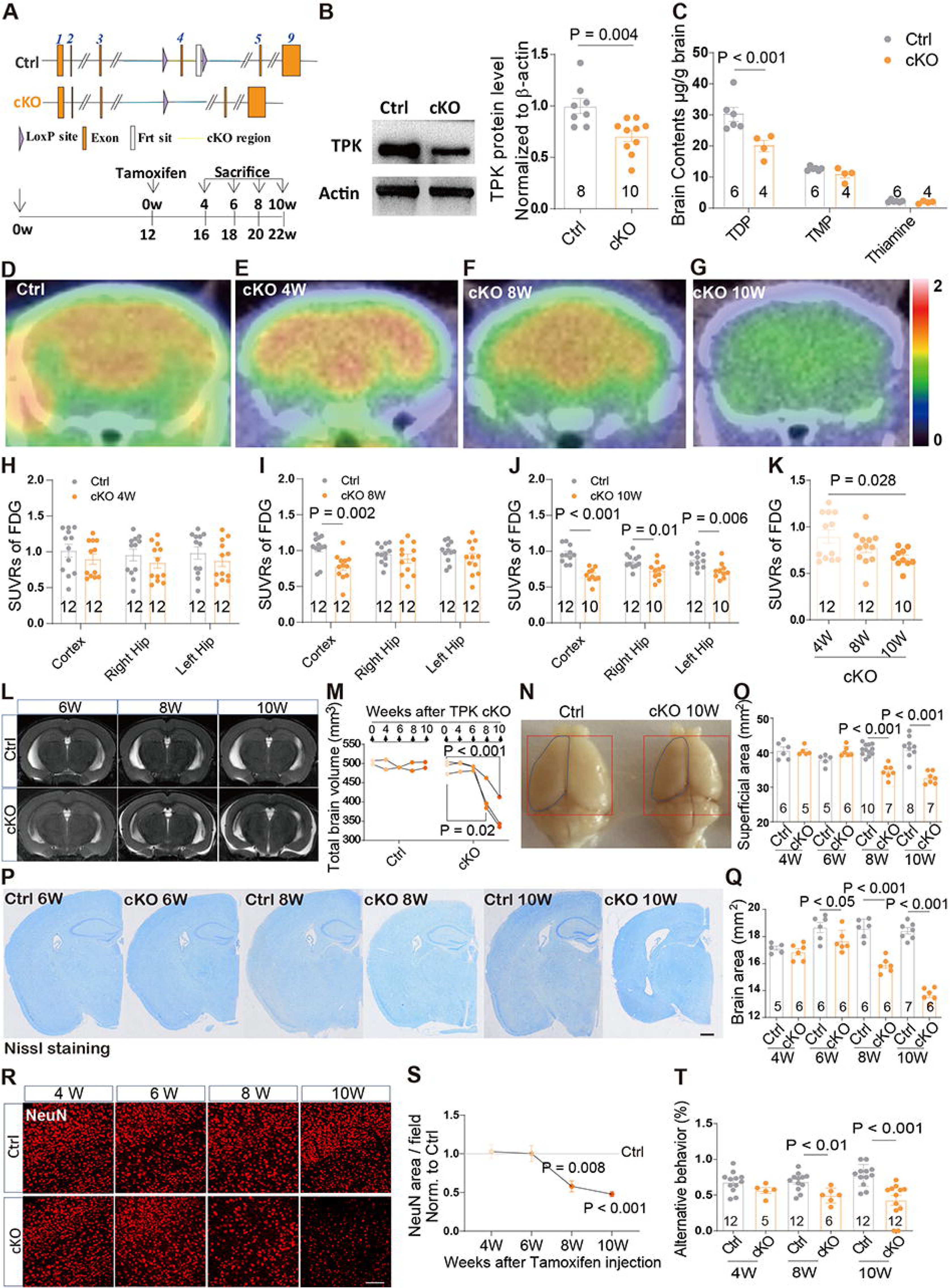
Mice with neuronal *Tpk* knockout manifest impairments in cerebral glucose metabolism, progressive loss of neurons, and brain atrophy. Panel A shows strategy of constructing the model with conditional knockout of *Tpk* in brain excitatory neurons of adult mice (*Tpk*-cKO mice). Panel A shows significant reduction of TPK protein levels in brain samples of the *Tpk*-cKO mice compared to their control littermates. Panel C shows the levels of TDP, but not that of thiamine monophosphate (TMP) and thiamine, in brain samples of the *Tpk*-cKO mice were significantly lower than that in their control littermates. Panel D-G shows representative images of brain FDG-PET / CT of the control littermates (D) and *Tpk*-cKO mice in the 4^th^ (E), 8^th^ (F), and 10^th^ (G) weeks after tamoxifen treatment. Panel H-J shows statistical results on the levels of cortical and hippocampal FDG uptake in the control littermates and *Tpk*- cKO mice in the 4^th^ (H), 8^th^ (I), and 10^th^ (J) weeks after tamoxifen treatment. Panel K shows cortical FDG uptake in the *Tpk*-cKO mice manifested a progressive decline in the 4^th^, 8^th^, and 10^th^ weeks after tamoxifen treatment. Panel L shows representative images of structural brain T2 weighted MRI of the control and *Tpk*-cKO mice in the 6^th^, 8^th^, and 10^th^ weeks after tamoxifen treatment. Panel M shows progressive declines of total brain volumes based on MRI quantification in individual *Tpk*-cKO mice (n = 3) in the 8^th^ and 10^th^, but not the 4^th^ and 6^th^, weeks after tamoxifen treatment as compared with the self-brain volumes before tamoxifen treatment. The brain volumes of their control littermates had no significant changes at all observation time points (n = 2). Panel N shows photographs of representative brain samples of the control littermates and *Tpk*-cKO mice in the 10^th^ week after tamoxifen treatment. Panel O shows quantification of brain superficial areas, as outlined in blue in (N), showing significant reduction in the *Tpk*-cKO mice compared to their control littermates in the 8^th^ and 10^th^, but not in the 4^th^ and 6^th^, weeks after tamoxifen treatment. Panel P and Q shows representative images (P) and quantifications of brain areas (Q) through Nissl staining. Significant decreases in brain areas were found in the *Tpk*-cKO mice compared to control littermates in the 6^th^, 8^th^, and 10^th^ weeks, but not in the 4^th^ week. Panel R and S shows representative images and quantifications of the areas of NeuN-positive cells in cortical regions. Significant decreases were found in the *Tpk*-cKO mice compared to control littermates in the 8^th^ and 10^th^ weeks, but not in the 4^th^ and 6^th^ weeks. Panel T shows significantly lower alternative ratios in Y maze test of the *Tpk*-cKO mice than their control littermates in the 8^th^ and 10^th^ weeks, but not in the 4^th^ week, after tamoxifen treatment. Summary data represent means ± SEM. Scale bars in panels (R), 100 µm; in panels (P), 500 µm. P values are indicated or > 0.05 if not labeled.

### Abnormal glucose metabolism in the *Tpk*-cKO mice

Brain glucose metabolism was first assayed using FDG-PET / CT in the 4^th^, 8^th^, and 10^th^ weeks after tamoxifen injection. Compared to control littermates, the *Tpk*-cKO mice exhibited a significant decrease in glucose uptake initially in the cortices in the 8^th^ week, which was then extended to more brain regions in the 10^th^ week (Fig. 3D-J; Fig. S6C-E). Self-comparison analysis showed a gradual decline of glucose uptake in the cortices of the *Tpk*-cKO mice (Fig. 3K). Moreover, the cortices of the *Tpk*-cKO mice in the 10^th^ week revealed significant accumulations of many substrates involved in glycolysis, especially glucose and glucose 6-phosphate, but decreased levels of those involved in oxidative phosphorylation, as indicated by metabonomic analysis (Fig. S7A-C). These results demonstrate that TPK deficiency significantly disrupts intracellular glucose metabolism and mitochondrial function, leading to a compensatory shift from the primarily aerobic oxidative phosphorylation pathway to glycolysis, which drastically impairs energy metabolism.

The *Tpk*-cKO mice exhibited a gradual increase in body weight from the 6^th^ week after tamoxifen treatment compared to control littermates (Fig. S7D), suggesting a change in metabolic status in the mutant mice. To test if neuronal TPK deficiency is also linked to peripheral glucose homeostasis, glucose tolerance test was performed in the 4^th^, 8^th^, and 10^th^ weeks after tamoxifen injection. In all three weeks, the *Tpk*-cKO mice manifested significant abnormality in glucose tolerance (Fig. S7E-H). Western blotting showed that the level of insulin receptor substrate-1 (IRS1) was significantly reduced, while the ratio of phosphorylated IRS-1 to total IRS-1 was significantly increased in brains of the *Tpk*-cKO mice in the 10^th^ week after tamoxifen treatment. The expression of IRβ was not affected, although the ratio of phosphorylated IRβ to total IRβ showed a tendency to decrease (Fig. S7I). Together, these results indicate that *Tpk* deficiency in brain excitatory neurons of adult mice impairs the homeostasis of cerebral and peripheral glucose metabolism, resulting in insulin resistance.

### Progressive loss of synapses and neurons causing brain atrophy and cognitive dysfunction in the *Tpk*-cKO mice

The brain volume of the *Tpk*-cKO mice exhibited a progressive decrease from the 8^th^ week, but not before the 6^th^ week, after tamoxifen treatment as detected longitudinally by magnetic resonance imaging (MRI; Fig. 3L, M). The result was supported by the measurement of brain superficial areas, total brain weights, cortical weights, and hippocampal weights (Fig. 3N, and Q; Fig. S8A-C). No significant changes were found in the cerebellum weight of the *Tpk*-cKO mice except for a moderate decrease in the 10^th^ week (Fig. S8D). Histological examination further demonstrated a progressive brain atrophy and loss of cerebral neurons in the *Tpk*-cKO mice beginning in the 8^th^ week after tamoxifen treatment (Fig. 3P-S; Fig. S8E-G), consistent with the results measured by structural (MRI) and volumetrics. The loss of cortical synapses in the *Tpk*-cKO mice was detectable in the 6^th^ week (Fig. S8H-L), earlier than the detectable loss of neurons.

A battery of behavioral tests was used to examine cognitive functions and other behaviors in the *Tpk*-cKO mice. Morris water maze tests showed that the *Tpk*-cKO mice had significant decreases in escape latency and crossing times in the 8^th^ week compared to the control mice and the *Tpk*-cKO mice in the 4^th^ week after tamoxifen treatment. The swimming speed of the *Tpk*-cKO mice in the 8^th^ week was also decreased (Fig. S9A-D). In addition, the *Tpk*-cKO mice also displayed decreases in the maximal speed and duration in the rotarod assay in the 8^th^ and 10^th^ weeks after the induction (Fig. S9E, F). As the reluctance of the *Tpk*-cKO mice to move could complicate the interpretation of results of the forced behavioral tests like the Morris water maze and rotarod assays, we performed the open-field and the Y-maze assays, in which the animals were not forced to move. The open-field assay revealed no significant differences in the distance traveled within 10 min, the average speed traveled, and the percentage of time spent in the center of the arena between the *Tpk*-cKO mice and their control littermates, suggesting no changes in spontaneous activity and anxiety levels (Fig. S9G-I). The Y-maze assay, however, showed that the *Tpk*-cKO mice had significant impairment in spatial working memory in the 8^th^ week after tamoxifen treatment (Fig. 3T).

### Aβ and Tau pathologies in the *Tpk*-cKO mice

ELISA was used to detect Aβ40 and Aβ42 levels in cortices of the control and *Tpk*-cKO mice. The total Aβ42 level was elevated in the *Tpk*-cKO mice from the 6^th^ week, while the total Aβ40 level did not enhance until the 10^th^ week after tamoxifen treatment. The insoluble Aβ42 level was significantly increased from the 6^th^ week, while the soluble Aβ42 level was increased from the 8^th^ week after tamoxifen induction. The insoluble Aβ40 level was not significantly increased, while the soluble Aβ40 level was increased in the 10^th^ week. The ratio of insoluble Aβ42 / Aβ40 was significantly increased in the 10^th^ week, while the ratio of soluble Aβ42 / Aβ40 was significantly increased in the 8^th^ week after tamoxifen treatment (Fig. 4A-C; Fig. S10A-I). Furthermore, the levels of Aβ precursor protein (APP), β-site APP cleaving enzyme 1 (BACE1), *C*-terminal fragment-α (CTFα), and *C*-terminal fragment-β (CTFβ) were significantly elevated in cortical samples of the *Tpk*-cKO mice in the 10^th^ week (Fig. 4D, E).

**Fig 4.**
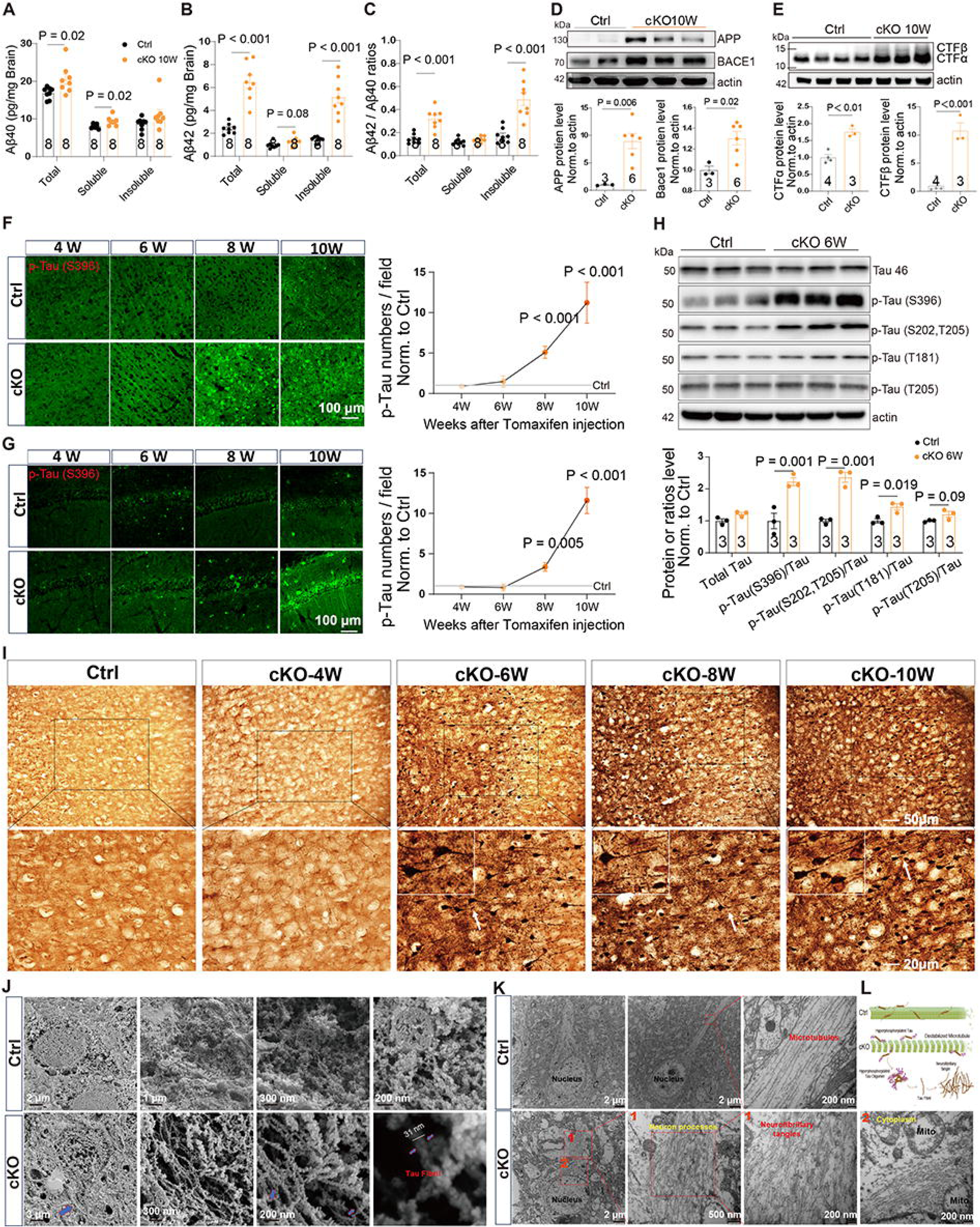
Brain Aβ and Tau pathologies in the *Tpk*-cKO mice. Panel A-C shows comparisons of the levels of total, soluble, and insoluble Aβ40 (A), Aβ42 (B), and Aβ42 / Aβ40 ratios (C) in cortical samples between the control littermates (Ctrl) and *Tpk*-cKO mice in the 10^th^ week after tamoxifen treatment. The levels of total Aβ40, total Aβ42, soluble Aβ40, insoluble Aβ42, and the ratios of total Aβ42 / total Aβ40 and insoluble Aβ42 / insoluble Aβ40 were significantly increased in the *Tpk*-cKO samples. Panel D shows images of Western blots and quantification showing that both APP and BACE1 proteins were significantly enhanced in cortical samples of the *Tpk*-cKO mice compared to their control littermates in the 10^th^ week after tamoxifen treatment. Panel E shows **i**mages of Western blots and quantification showing that both CTFα and CTFβ proteins were significantly elevated in cortical samples of the *Tpk*-cKO mice compared to their control littermates in the 10^th^ week after tamoxifen treatment. Panel F shows images and quantification of phosphorylated Tau (p-Tau 396), as revealed by immunofluorescent staining using the phospho-S396 antibody, showing that Tau phosphorylation was significantly increased in the *Tpk*-cKO mice compared to their control littermates in the 8^th^ (n = 7, 5, for Ctrl and cKO, respectively, and the same convention herein, P < 0.001) and 10^th^ weeks (n = 6, 4, P < 0.001), but not in the 4^th^ and 6^th^ weeks (n = 6, 6, P > 0.05) after tamoxifen treatment. Panel G shows similar to F but for hippocampal samples showing no significant changes in the *Tpk*-cKO mice compared to their control littermates in the 4^th^ and 6^th^ weeks (n = 6, 6, P > 0.05 for both weeks), while significant increases in Tau phosphorylation in the 8^th^ (n = 7, 5; P = 0.005) and 10^th^ (n = 6, 4; P < 0.001) weeks after tamoxifen treatment. Scale bars for (F) and (G), 50 µm. Panel H shows Images of Western blots and quantification of total Tau and p-Tau levels as well as the p-Tau / Tau ratios showing significantly enhanced phosphorylation at multiple sites in Tau in cortical tissues of the *Tpk*-cKO mice at the 6^th^ week after tamoxifen treatment compared to their control littermates. Panel I shows representative images of Bielschowsky silver staining showing the presence of neurofibrillary tangles in cortical samples of the *Tpk*-cKO mice starting from the 6^th^ week after tamoxifen treatment. Scale bars, 50 µm in the upper row and 20 µm in the lower row. Panel J shows scanning electron microscopic images of neurofibrillary tangles, with diameters of approximately 31 nm, in the cytoplasm of cortical neurons in the *Tpk*-cKO mice in the 10^th^ week after tamoxifen treatment. Neurofibrillary tangles were not found in neurons of the control littermates. Panel K shows neurofibrillary tangles detected by transmission electron microscopy in the processes (Area 1) and cytoplasm (Area 2) of neurons of the *Tpk*-cKO mice in the 10^th^ week after tamoxifen treatment. Panel L shows the schematic diagram of neurofibrillary tangles formation in the *Tpk*-cKO mice. Scale bars are indicated in the images. Mito, mitochondrion. P values are indicated above the data points and bars or > 0.05 if not labeled.

Increased density of paired helical filaments (PHF)-Tau stained by the anti-phospho Tau (S396) antibody was observed in both cortical and hippocampal tissues of the *Tpk*-cKO mice beginning in the 8^th^ week and becoming more pronounced in the 10^th^ week after tamoxifen treatment (Fig. 4F, G). The hyperphosphorylated Tau was found primarily in neurons (Fig. S11A). Western blotting revealed that Tau was hyperphosphorylated at multiple sites (S396, S202, T205, T181) in cortical tissues of the *Tpk*-cKO mice in the 6^th^ week after tamoxifen treatment (Fig. 4H). Using two different silver staining methods, Bielschowsky ^34^ and Gallyas ^35, 36^, we further demonstrated the presence of neurofibrillary tangles in brain neurons of the *Tpk*-cKO mice beginning in the 6^th^ week and becoming more pronounced in the 8^th^ and 10^th^ weeks after tamoxifen treatment, which were absent in that of the control littermates (Fig. 4I; Fig. S11B). Scanning electron microscopy analysis visually showed neurofibrillary tangles in neuronal cytoplasm, with a diameter of approximately 31 nm. Transmission electron microscopy also revealed clear neurofibrillary tangles in brain neurons of the *Tpk*-cKO mice (Fig. 4J-L).

### Glial activation and neuroinflammation in the *Tpk*-cKO mice

The *Tpk*-cKO mice showed significant activation of microglia in cortical tissues from the 4^th^ week after tamoxifen treatment (Fig. S12A-C), representing one of the earliest changes detected in the brains of the mutant mice. Microglial activation was detected in the hippocampus beginning from the 6^th^ week (Fig. S12D-F). In addition, astrocytes were significantly activated in cortices from the 4^th^ week and in the hippocampus from the 8^th^ week after the induction (Fig. S12G-L). In line with glial activation, the levels of tumor necrosis factor-α (TNF-α) mRNA in cortical samples of the *Tpk*-cKO mice were significantly enhanced starting from the 4^th^ week after tamoxifen treatment, while the mRNA levels of other inflammatory factors, such as interleukin-6 and interleukin-1β, exhibited significant increases beginning from the 6^th^ week (Fig. S12M-O).

### Therapeutic effects of boosting thiamine metabolism on AD-like phenotypes induced by *Tpk* deficiency

The recently developed adeno-associated virus (AAV)-CAP-B10 vector can cross the blood-brain barrier and preferentially target neurons ^37^. To test the gene therapeutic potential of targeting TPK, AAV-CAP-B10 encoding *Tpk* (AAV-TPK) was injected to restore *Tpk* expression in the *Tpk*-cKO mice at the 5^th^ week after tamoxifen treatment. Robust increase in TPK protein level was detected in both the control and *Tpk*-cKO mice at the 4^th^ week after AAV-TPK injection (Fig. S13A-D). The results from Y-maze, nest-building, and rotarod behavior tests showed that TPK re-expression significantly ameliorated cognitive impairment and motor disorder of the *Tpk*-cKO mice (Fig. 5A, B; Fig. S13E, F). Consistently, the neuronal loss and brain atrophy of the *Tpk*-cKO mice were rescued by the restoration of TPK expression (Fig. 5C-E; Fig. S13G-I). Also, the enhancements of Aβ, APP, BACE1, Tau phosphorylation, and glial activation in the *Tpk*-cKO mice were significantly suppressed (Fig. 5F-H). On the other hand, the overexpression of TPK had little effects on behaviors, brain Aβ and phospho-Tau levels, and glial activation in the wild-type mice (Fig. S13).

**Fig 5.**
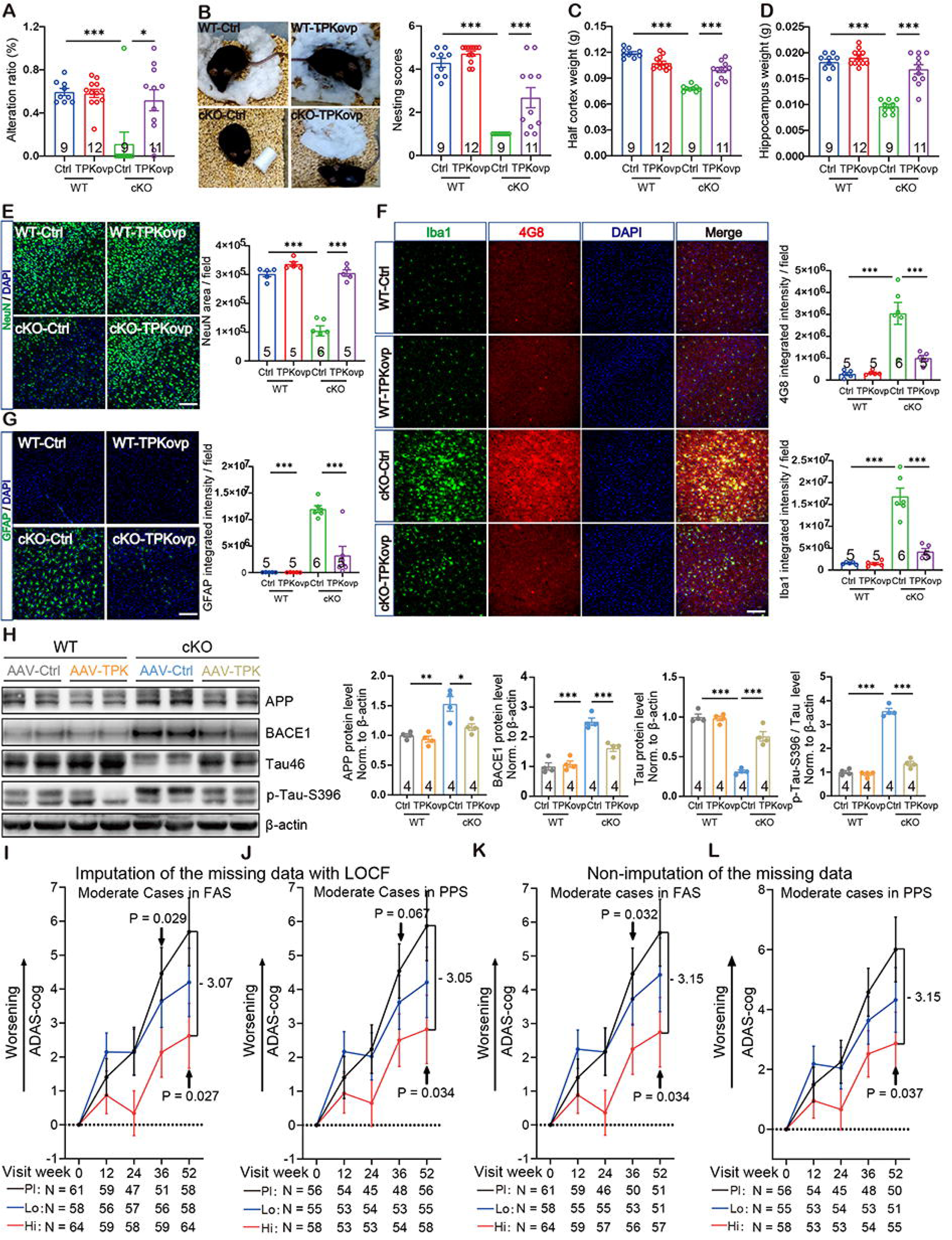
Alleviation of the deficits of *Tpk*-cKO mice by restoration of TPK and attenuation of cognitive decline of AD patients by benfotiamine treatment. Panel A shows restoration of TPK in the brain rescued the decreased alternative ratio in Y-maze test of the *Tpk*-cKO mice. Panel B shows nest-building assessed after 24 hours of housing was impaired in the *Tpk*-cKO mice compared to their control littermates, which was rescued by restoration of TPK in the brain. Panel C and D show the decreased cortical (C), and hippocampal (D) weights of the *Tpk*-cKO mice were rescued by restoration of TPK in the brain. Panel E-G show the representative images and quantification of immunohistochemical staining of cortical sections by NeuN (E), 4G8 and Iba1 (F), and GFAP (G) antibodies, showing that the neuronal loss, Aβ enhancement, and microglial and astrocyte activation in the *Tpk*-cKO mice were rescued by restoration of TPK in the brain. Panel H shows images of Western blots and quantification of APP, BACE1, Tau, and p–Tau (S396) showing that the enhancements in the levels of APP and BACE1 proteins and the ratio of p-Tau / Tau, as well as the reduction in the total Tau level, in the *Tpk*-cKO mice were rescued by restoration of TPK in the brain. Summary data represent means ± SEM. Scale bars, 100 µm. P values are indicated above the bars for *P < 0.05, **P < 0.01, ***P < 0.001 or > 0.05 if not labeled. Panel I and J shows the mean changes of ADAS-cog scores at the indicated time points from the baseline in moderate AD cases defined by the MMSE scores of 11 to 19 at enrollment among high-dose benfotiamine (600 mg daily, Hi), low-dose benfotiamine (300 mg daily, Lo), and placebo (Pl) groups in full analysis set (FAS) and per protocol set (PPS). The enhancement of ADAS-cog scores from baseline was significantly less in the high-dose group than that in placebo group at the 52^nd^ week in both FAS (I) and PPS (J) by MMRM analyses. The missing data of the ADAS-cog score were imputed using the last observation carried forward (LOCF) method. Panel K and L shows MMRM analysis of the same data as in (I) and (J) without imputation of the missing ADAS-cog scores. The enhancement of ADAS-cog scores from baseline was significantly less in the high-dose group than that in placebo group at the 52^nd^ week in both FAS (K) and PPS (L). Bars indicate standard deviation. N indicates the number of patients in each group or subgroup at the tested time point. Three cases in the low-dose benfotiamine group were not included in the analyses because their MMSE scores deteriorated from 11 at enrollment to 10 at baseline.

To further explore whether TPK exerts its effects through its product TDP or not, N2a cells with stable *Tpk* knockdown using RNAi were utilized. These cells displayed a decrease in TPK protein level to approximately 40% of that in control N2a cells (Fig. S14A) and concomitant increases in the levels of APP and BACE1 proteins (Fig. S14B). Not only did overexpression of HA-tagged TPK restore APP and BACE1 proteins to control levels (Fig. S14B), but also the supplementation of thiamine and TDP significantly decreased the APP and BACE1 levels in the *Tpk* knockdown N2a cells (Fig. S14C, D). Together, the above results suggest that the reduced production of TDP due to TPK deficiency underscores the pathophysiological features observed in both the *Tpk*-cKO mice.

### Benfotiamine dose-dependently delays cognitive decline of AD patients

A total of 434 patients were screened and 302 were randomly assigned to placebo (n = 99), low-dose benfotiamine (300 mg/day, n = 104), and high-dose benfotiamine (600 mg/day, n = 99) groups. The demographic data and clinical characteristics at the baseline are listed in Table S3. For analysis, the full-analysis set (FAS), in which the subjects received at least one post-baseline efficacy assessment, included 97 in the placebo (98.0%), 98 in the low-dose (94.2%), and 97 in the high-dose groups (98.0%), while the per-protocol set (PPS), in which the subjects fulfilled the protocol in term of the efficacy assessment, had 89 in the placebo (89.9%), 86 in the low-dose (82.7%), and 90 in the high-dose groups (90.9%, Fig. S15).

Based on the MMSE scores at enrollment, the patients had either mild AD (MMSE scores 20 - 24) or moderate AD (MMSE scores 11 – 19, Table S3). When analyzed together without stratification (overall population analysis), the increase in ADAS-cog scores showed a trend to slow down by benfotiamine in a dose dependent manner over the 52 weeks period. However, the data did not reach statistical significance, as analyzed using the mixed model for repeated measures (MMRM), no matter if the missing data were imputed or not by the last observation carried forward (LOCF) method (Fig. S16).

Noticeably, the ADAS-cog scores of the placebo group varied greatly at the primary endpoint of week 52, with the majority of the mild patients (20 ≤ MMSE scores ≤ 24, at the time of enrollment) exhibiting no obvious increase at all or even decreases (Fig. S17). This may be attributed to the inclusion of donepezil (5 mg daily) as the base medication to all patients in this trial. Obviously, if the mild AD did not have detectable cognitive decline during the trial period, then it would be hard for the test drug to show any benefit. Therefore, we removed mild cases from the analysis. The remaining cases (moderate AD with MMSE scores 11 – 19 at enrollment) accounted for 59 – 66% of the total patients in individual treatment groups (Table S3), and to them, benfotiamine exhibited a clear benefit against cognitive deterioration, with dose and time dependence. At the primary endpoint of week 52, the mean changes of ADAS-cog scores in the high-dose subgroup (2.62, 95% CI 0.74 to 4.50 in FAS; 2.82, 95% CI 0.85 to 4.79 in PPS. The same below) was significantly less than that in the placebo subgroup (5.69, 95% CI 3.73 to 7.64; 5.87, 95% CI 3.86 to 7.88; Net difference: -3.07, -3.05; p = 0.027, 0.034 for the missing data imputed using LOCF). The low-dose subgroup exhibited a trend to suppress the increase of ADAS-cog scores (4.20, 95% CI 2.22 to 6.18; 4.21, 95% CI 2.18 to 6.24; Net difference: -1.49, -1.66; p = 0.292, 0.253), which did not reach statistical significance. Interestingly, at weeks 12 and 24, the effect of high-dose benfotiamine was not significant, but at week 36, it began to show a significant effect in preventing cognitive impairments compared to placebo (Net difference: -2.32, -2.04; p = 0.029, 0.067, Fig. 5I, J). Similar results were also obtained without imputation of the missing data (Fig. 5K, L). Together, the above data demonstrate that benfotiamine dose-dependently suppresses cognitive decline of patients with moderate AD.

## Discussion

The distinct pathophysiological features and clinical presentations of each disease necessitate that the key contributor(s) identified in the pathogenesis of each disease should be disease-specific. Our studies showed that TPK down-regulation was specific for AD because the other genes associated with thiamine metabolism did not exhibit significant expression inhibition in AD and the expressions of all four genes also had not significant changes in other neurodegenerative diseases. High TPK expression is protective against AD pathology, as indicated by the positive correlation trend between TPK mRNA levels and the scores of Braak staging in control NC subjects but a significant negative correlation in AD patients, as well as the significantly higher TPK mRNA level in NC controls than AD patients with cognitive impairment under the condition of the same Braak stage □ (Fig. 1; Table S1). The results imply that TPK inhibition contributes to pathophysiological alterations and cognitive decline of AD.

In order to demonstrate whether brain Aβ deposition and diabetes, a well-known risk factor of sporadic AD ^20, 21^, cause the disease through TPK inhibition, classical mouse and cellular models of AD and diabetes were utilized. The results showed that the level of TPK protein in neurons close to the Aβ plaques was significantly reduced in the APP / PS1 transgenic mice (Fig. 2A). Oligomeric Aβ is considered as a main contributor of synaptic and neuronal impairment in AD because of its neurotoxicity ^38^. Further, oligomeric Aβ dose-dependently inhibited TPK expression in neurons cultured *in vitro* (Fig. 2B). Importantly, the reduction of TPK expression in neurons leaded to the enhancement of death and neurodegeneration biomarkers, which could be blocked by TPK re-expression (Fig. S3). High glucose, the synonymous with diabetes, selectively inhibited TPK expression in neurons cultured *in vitro*, but not in non-neuronal HEK 293 cells. The studies on the diabetic mice induced by STZ also showed that high glucose significantly inhibited the expression of TPK in the brains, but not in the livers. The results indicate that the inhibition of TPK expression caused by high glucose is selective for cell and tissue types.

The studies on the model of *Tpk* gene conditionally ablated in brain excitatory neurons of adult mice further show the decisive role in leading to multiple pathophysiological features of AD. The *Tpk*-cKO mice recapitulated all important multiple pathophysiological features of AD, including progressive loss of synapses and neurons causing brain atrophy and cognitive decline (Fig. 3), Aβ deposition and plaques, Tau hyperphosphorylation and neurofibrillary tangles (Fig. 4), brain glucose hypometabolism and deregulation of peripheral glucose metabolism (Fig. 3), and glial activation and neuroinflammation (Fig. S12). The *Tpk*-cKO mice represent the first demonstration that manipulation of a single AD-related gene can reproduce all major pathophysiological features of human AD in a mouse model, a dilemma previously thought to be impossible due to the significantly short lifespan and genetic differences between mice and humans. Our findings not only contradict these presumptions but also present a compelling case for their reconsideration.

Causative treatment targeting pathogenic factor is the most effective strategy to combat a disease. The restoration of *Tpk* expression in brains of the *Tpk*-cKO mice significantly attenuated the development of main pathophysiological features of the *Tpk*-cKO mice, including Aβ accumulation, Tau hyperphosphorylation, brain atrophy, and subsequent cognitive impairment (Fig. 5A-H). The supplementation of thiamine and TDP in the culture medium also abolished the increases of APP and BACE1 protein levels in N2a cells induced by *Tpk* knockdown (Fig. S14). These results indicate that targeting TPK and / or TDP deficiency has therapeutic potential in preventing the progression of multiple pathophysiological alterations of AD.

Clinical trial results of benfotiamine support the idea that TPK inhibition and hence TDP reduction can be an effective target for AD. Benfotiamine exhibited beneficial effects in delaying cognitive decline of early AD patients in a phase 2a clinical trial, with a reduction of deterioration by 77% based on the changes in Clinical Dementia Rating scale, compared to placebo at the one-year endpoint ^39^. The results of our current clinical trial further demonstrate significant efficacy in delaying cognitive decline of moderate AD patients (Fig. 5I-L). Further large-scale clinical trials are underway ^40^.

In summary, we show that brain TPK expression is specifically reduced in AD patients. Aβ deposition and diabetes also induce the inhibition of TPK expression in neurons. Importantly, conditional *Tpk* knockout in brain excitatory neurons of adult mice recapitulates all essential pathophysiological features of human AD in the symptomatic stage, testifying the critical role of TPK deficiency and hence TDP reduction in determining Aβ deposition and high glucose to multiple pathophysiological features of AD transition. The therapeutic effects of *Tpk* re-expression and thiamine and / or TDP supplement on cell and mouse models and AD patients also indicate a decisive role of TPK insufficiency and hence TDP reduction in neurons in deriving the transmission of multiple pathophysiological alterations of symptomatic stage for AD. Our findings bring novel perspectives and insights into AD research. Future studies should further investigate the role of TPK expression in the occurrence and development of Aβ pathology at preclinical stage of AD, so as to determine whether TPK inhibition and hence TDP reduction are a key pathogenic factor for AD.

## Data Availability

All data produced in the present study are available upon reasonable request to the authors.

## Acknowledgements

We thank Dr. Min Jiang and Molecular and Cellular Imaging Facility of Institutes of Brain Science (IOBS), Fudan University, for their support in confocal microscopy and data analysis, Dr. Qian Huang and Animal Behavior Facility of IOBS, Fudan University, for their support in animal behavioral experiments, and Dr. Yu Kong and Electron microscopy platform of CAS Center for Excellence in Brain Science and Intelligence Technology, State Key Laboratory of Neuroscience, Chinese Academy of Sciences, for their support in electron microscopic analysis.

## Author contributions

Sang, S. was responsible for the experimental procedures and data analysis from international datasets. Sang, S., Qian, T., Xia, Y., Zeng Y., Zhao, X., Cai, W., Jin, B., Qiu, H., Xu, Y., Huang, Q., Wang, C., and Cheng, X. were responsible for the pathological and rescue experiments of Tpk-cKO mice. Zhao, X., Zeng Y., Cai, W., were responsible for studying the effect of high glucose and Aβ oligomers on TPK protein expression. Peng, Y., Jin, B., and Cheng, X. were responsible for studying TPK expression in neurons of APP/PS1 transgenic mice. Cai, F., Zhang, Y., Zhang, Q., and Song, W. were responsible for studying the levels of TPK protein in brains of AD and control subjects. Jiang, D. and Guan, Y. were responsible for FDG-PET / CT. Wu, Y., Tong, H., and Zhong, K. were responsible for MR imaging. Huang, S. assisted animal model raising. Pan X., Zhao Q., Sang, S., and the benfotiamine phase 2 trial investigators were responsible for the clinical trial, which was initiated by Shanghai Raising Pharmaceutical Co., Ltd (Formerly known as Shanghai Rixin Biological Technology Co., Ltd.). Sang, S. was responsible for the verification and statistical analyses of the results. Yu, X. and Zhu, M.X. advised on animal experiments. Peng, Y., Yu, X., Zhu. M.X., and Song, W. revised the manuscript. Zhong, C. and Sang, S. designed the study and wrote the manuscript.

