## Supplementary figures and tables for "Thiamine pyrophosphokinase-1 deficiency in neurons drives Alzheimer’s multiple pathophysiological alterations"

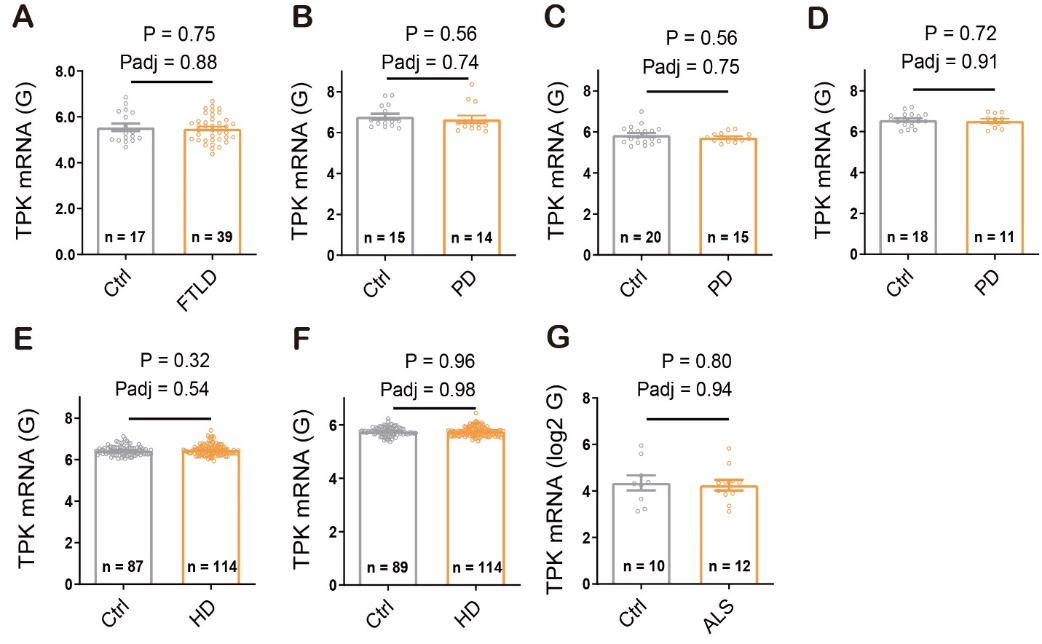

**Figure S1.** ***TPK* mRNA levels have no significant changes in brain samples of patients with non-AD neurodegenerative diseases.** Panel A shows no significant differences in *TPK* mRNA levels of frontal cortex samples between control subjects and patients with frontotemporal lobar degeneration (FTLD). Panels B-D show no significant differences in *TPK* mRNA levels between control subjects and patients with Parkinson’s disease (PD) in the GSE20168 dataset measuring prefrontal cortex samples (Panel B), in the GSE20291 dataset measuring putamen samples (Panel C), and in the GSE20292 dataset measuring substantia nigra samples (Panel D). Panels E and F show no significant differences in *TPK* mRNA levels between control subjects and patients with Huntington’s disease (HD) in the GSE3790A dataset (Panel E) and GSE3790B dataset (Panel F), both measuring cerebellum, frontal cortex, and caudate nucleus samples in two separate arrays. Panel G shows no significant differences in *TPK* mRNA levels of lumbar spinal cord samples between control subjects and patients with amyotrophic lateral sclerosis (ALS). Summary data represent means ± SEM. The number of subjects per group (n) is shown. P values are indicated above the bars. Adjusted P values (Padj) are determined using Benjamini-Hochberg method.

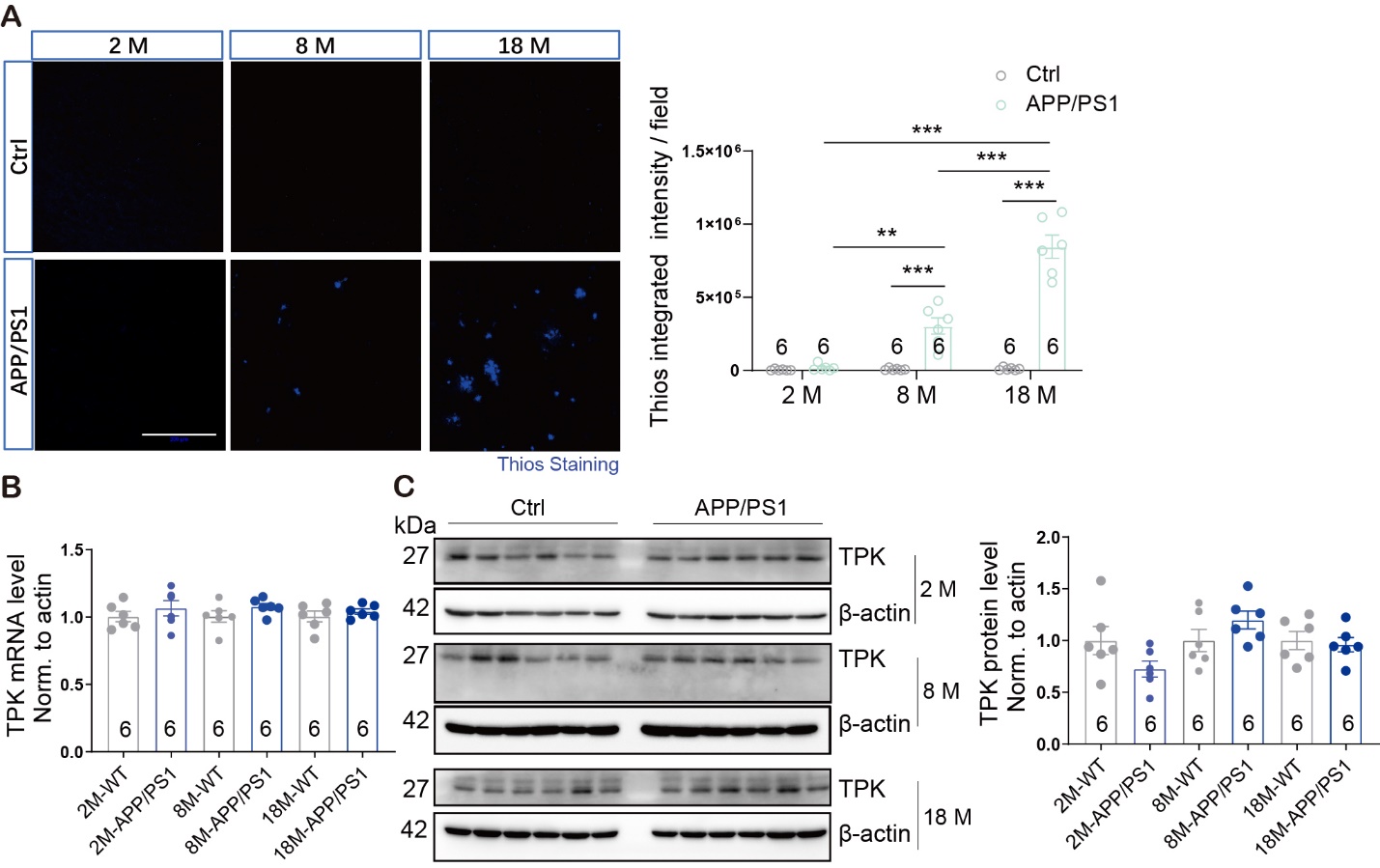

**Figure S2. No significant changes in TPK expression of cortical samples from the APP / PS1 transgenic mice.** Panel A shows representative images and quantifications of the integrated intensities of Aβ plaques using Thios staining in cortices of APP/PS1 transgenic mice. Significantly increased Aβ plaques were found in the APP/PS1 transgenic mice compared to control littermates in the 8- and 18-months-old mice, but not in the 2-months-old mice. Panel B shows no significant changes in the *Tpk* mRNA levels in the cortical samples of APP/PS1 transgenic mice compared to age-matched control mice, respectively. Panel C shows Western blot images and quantification of TPK protein in cortical tissues of APP/PS1 transgenic mice, showing that the level of TPK protein had no significant changes as compared with those in age-matched control mice, respectively. Scale bars, 200 µm. Summary data represent means ± SEM. The number of mice per group or repeated experiments is indicated in the bar. P values are indicated above the bars for *P < 0.05, **P < 0.01, ***P < 0.001 or > 0.05 if not labeled.

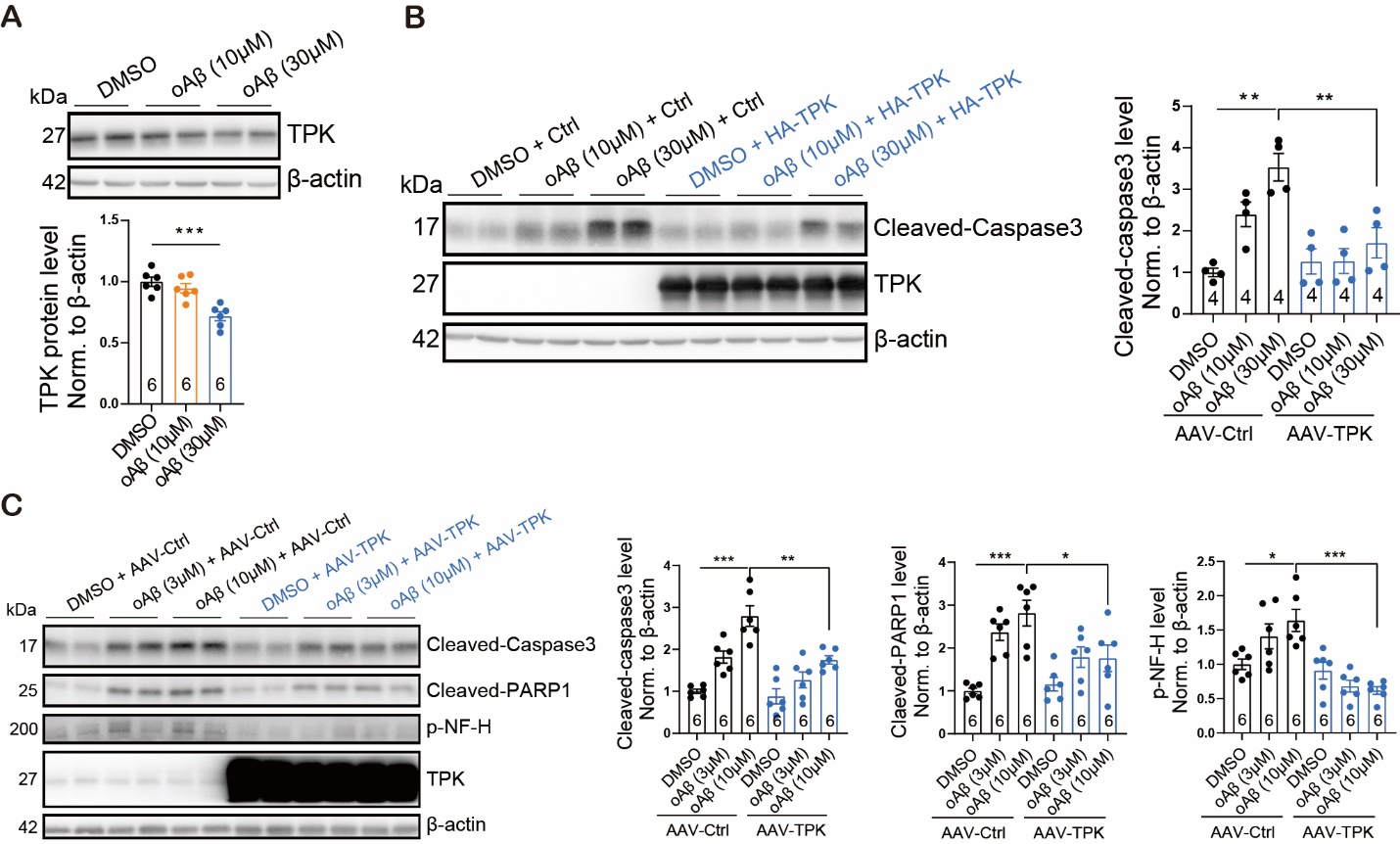

**Figure S3. TPK inhibition contributes to the neurotoxicity of oligomeric Aβ.** Panel A shows representative Western blot images and quantification of TPK protein, indicating that oligomeric Aβ significantly decreased the level of TPK protein in N2a cells. Panel B shows representative Western blot images and quantification of Cleaved-caspase3, indicating that the elevation of Cleaved-caspase3 proteins in N2a cells treated by oligomeric Aβ was rescued by *Tpk* overexpression. Panel C shows Western blot images and quantification of Cleaved-caspase3, Cleaved-PARP1, and p-NF-H, indicating that the elevation of those proteins in primary neurons treated by oligomeric Aβ was rescued by *Tpk* overexpression. Summary data represent means ± SEM. The number of mice per group or repeated experiments is indicated in the bar. P values are indicated above the bars for *P < 0.05, **P < 0.01, ***P < 0.001 or > 0.05 if not labeled.

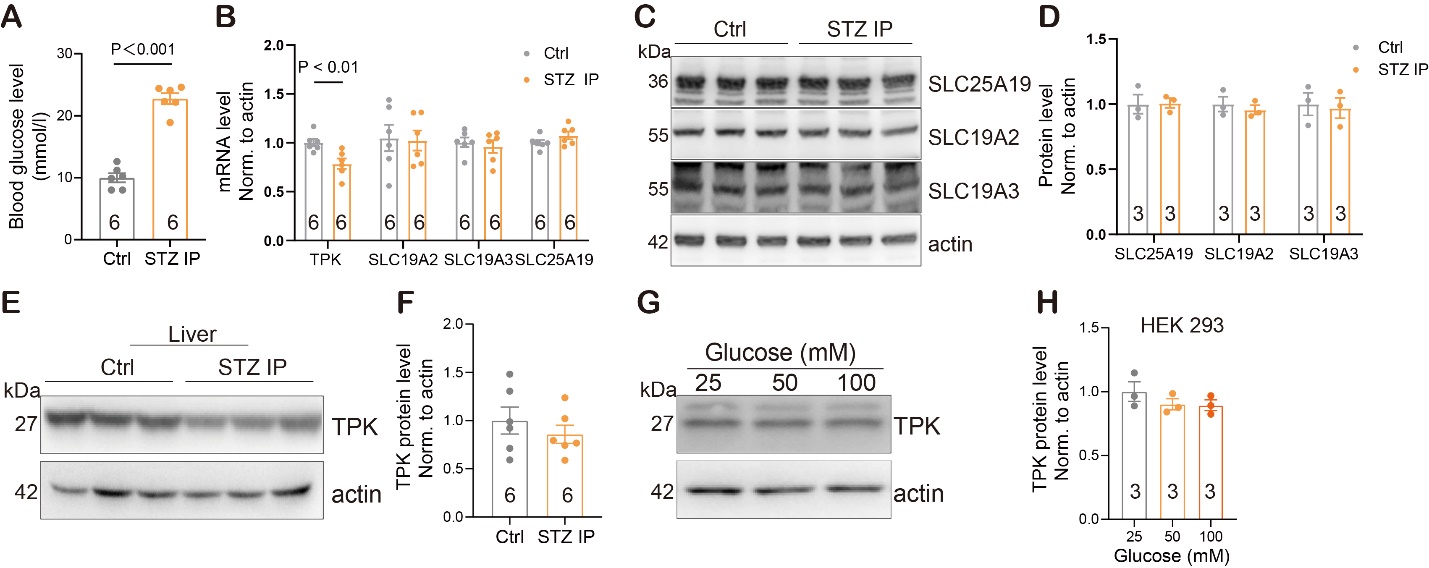

**Figure S4.** **Effects of STZ and high glucose treatment on the expression of genes associated with thiamine metabolism.** Panel A shows that the level of fasting blood glucose was significantly increased in mice 3 days after intraperitoneal STZ injection (STZ IP, 200 mg / kg). Panel B shows that among the four known genes associated with thiamine metabolism, only *Tpk* mRNA level was significantly decreased in brain samples of mice treated by STZ mice as compared with that in control mice treated by solvent. The mRNA levels of *Slc19a2*, *Slc19a3*, and *Slc25a19* were not significantly altered. Panel C and D shows representative Western blot images and quantification of SLC19A2, SLC19A3, and SLC25A19 proteins in brain tissues of mice with solvent (Ctrl) or STZ injection, showing no significant changes between the two groups. Panel E and F shows representative Western blot images and quantification of TPK protein in liver tissues of mice with solvent or STZ injection, showing no significant changes between the two groups. Panel G and H shows representative Western blot images and quantification of TPK protein in HEK293 cells, showing no significant reduction in the level of TPK protein after high glucose treatment compared to normal glucose (25 mM) treatment. Summary data represent means ± SEM. The number of mice per group or repeated experiments is indicated in the bar. P values are indicated above the bars or > 0.05 if not labeled.

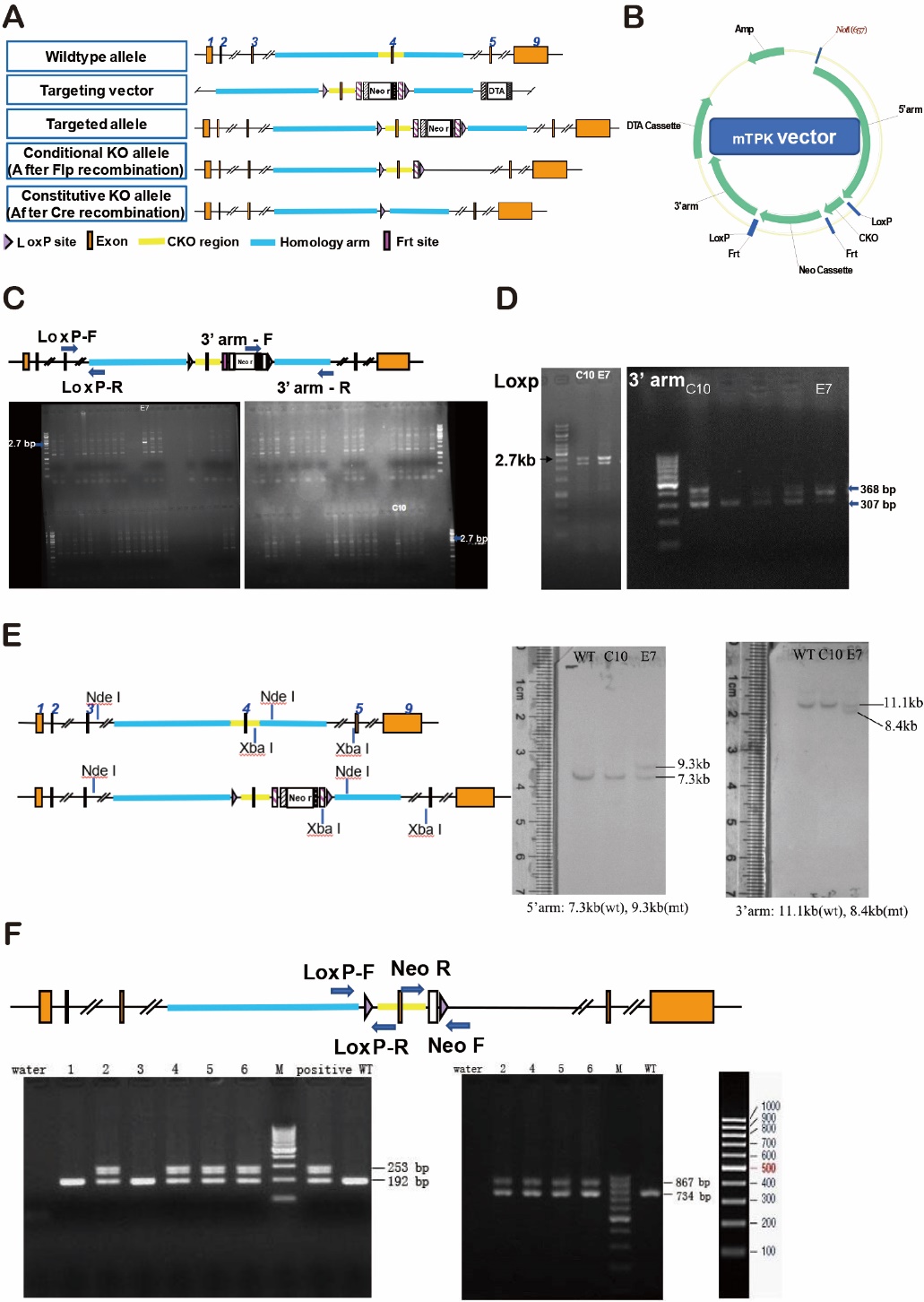

**Figure S5. Establishment of the model with conditional *Tpk* knockout in cerebral excitatory neurons of adult mice.** Panel A shows the overview of the targeting strategy. In the targeting vector, the Neo cassette was flanked by Frt sites and the cKO region flanked by LoxP sites. Panel B showed the map of the targeting vector, with DTA used for negative selection. Panel C showed that 2 positive clones, C10 and E7, were identified from 85 clones screened with 3’ arm F / 3’ arm R primers for a ~2700-bp product. Re-confirmation of the targeted clones with 3’ arm F / 3’ arm R primers showing the ~2700-bp fragment in C10 and E7. Panel D showed further verification of the clones with LOXP-F / LOXP-R primers for a 307-bp fragment from the wild-type allele and a 368-bp fragment from the recombinant allele. Panel E showed Southern Blot analysis conducted by 5’-probe and 3’-probe. For 5’-probe, genomic DNAs of clones C10 and E7 were digested by *Nde* I and analyzed by Southern blotting for a 7.3-kb band from wild type allele and a 9.3-kb band from recombinant allele; E7 was found positive. For 3’-probe, genomic DNAs of clones C10 and E7 were digested by *Xba* I and analyzed by Southern blotting for an 11.1-kb band from wild type allele and an 8.4-kb band from recombinant allele. E7 was found positive. Panel F showed PCR identification of the F1 mice. Out of 6 pups, 4 (pups 2, 4, 5, 6 from clone E7) were identified to be positive by the first PCR screening with primers LoxP_F / LoxP_R with the expected 192-bp fragment from wild type allele and 253-bp fragment from recombinant allele (left gel). The tails of the 4 positive pups were recut for the second PCR reconfirmation with a different pair of primers Neo _F / Neo _R and all of them yielded the 867-bp fragment expected from recombinant allele, in addition to the 734-bp fragment from the wild type allele.

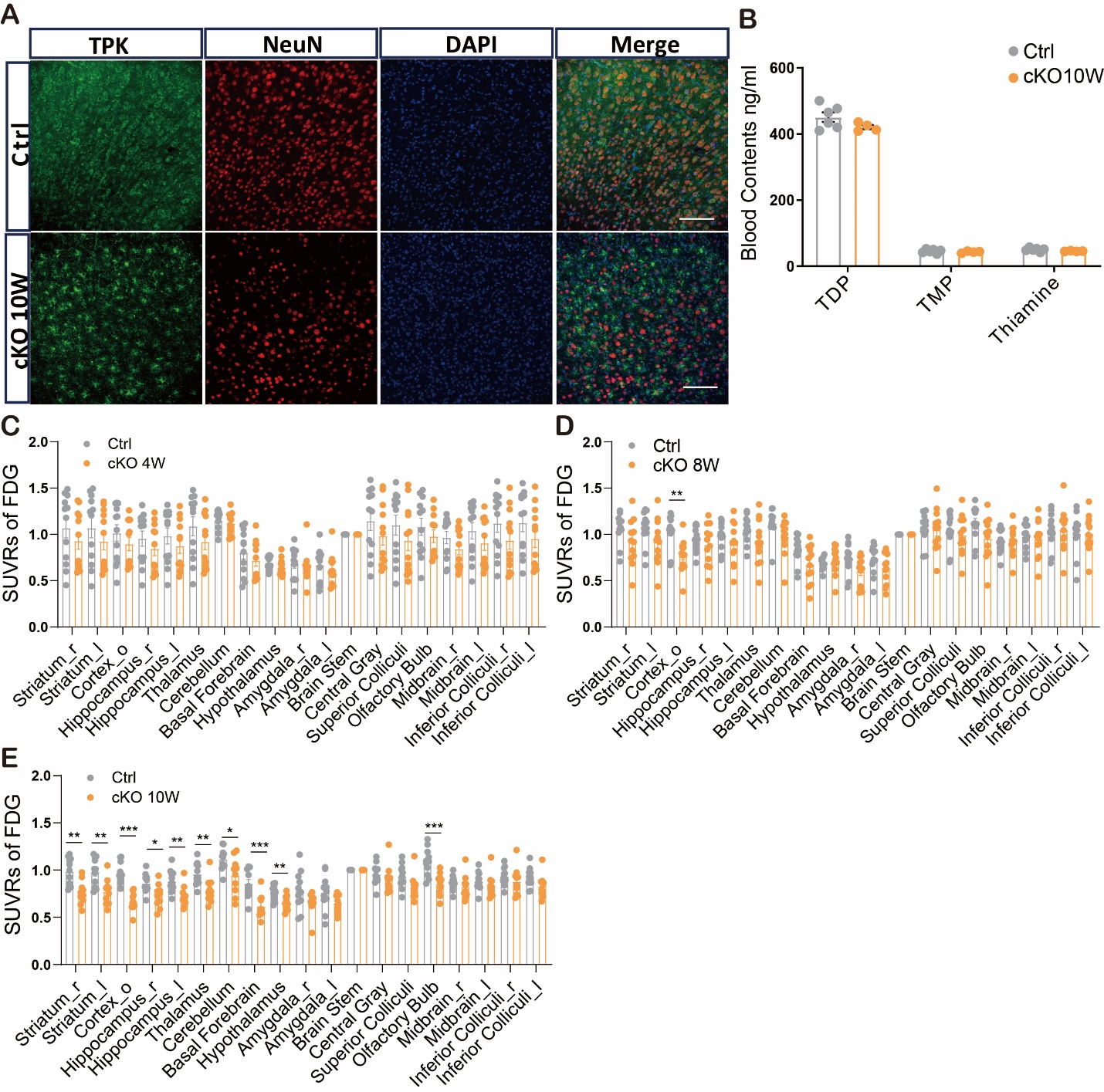

**Figure S6. Selective loss of TPK protein in brain neurons of the *Tpk*-cKO mice decreased the level of FDG uptake in different brain regions without affecting the levels of thiamine and its phosphate esters in the blood.** Panel A shows the representative images of immunohistochemical staining, showing that the co-existence of TPK protein with neurons stained by the NeuN antibody was significantly reduced in cortical samples of the *Tpk*-cKO mice in the 10^th^ week after tamoxifen treatment as compared with that in control littermates (Ctrl). Scale bars, 100 µm. Panel B shows no significant differences in the levels of blood TDP, thiamine monophosphate (TMP), and thiamine between the control littermates (Ctrl, n = 4) and *Tpk*-cKO mice (n = 6, all P values > 0.05) in the 10^th^ week after tamoxifen treatment. Panel C shows no significant differences in FDG uptakes in all tested brain regions between the control littermates and *Tpk*-cKO mice in the 4^th^ week after tamoxifen treatment (n = 12, P > 0.05). Panel D shows a significant decrease appeared in the cortex (P = 0.002), but not in other brain regions, of the *Tpk*-cKO mice compared to their control littermates in the 8^th^ week after tamoxifen treatment (n = 12, P > 0.05). Panel D shows significant decreases were detected in the cortex, hippocampus, striatum, thalamus, cerebellum, basal forebrain, hypothalamus, and olfactory bulb of the *Tpk*-cKO mice (n = 10) compared to their control littermates in the 10^th^ week after tamoxifen treatment (n = 10). *P < 0.05, **P < 0.01, ***P < 0.001 or > 0.05 if not labeled.

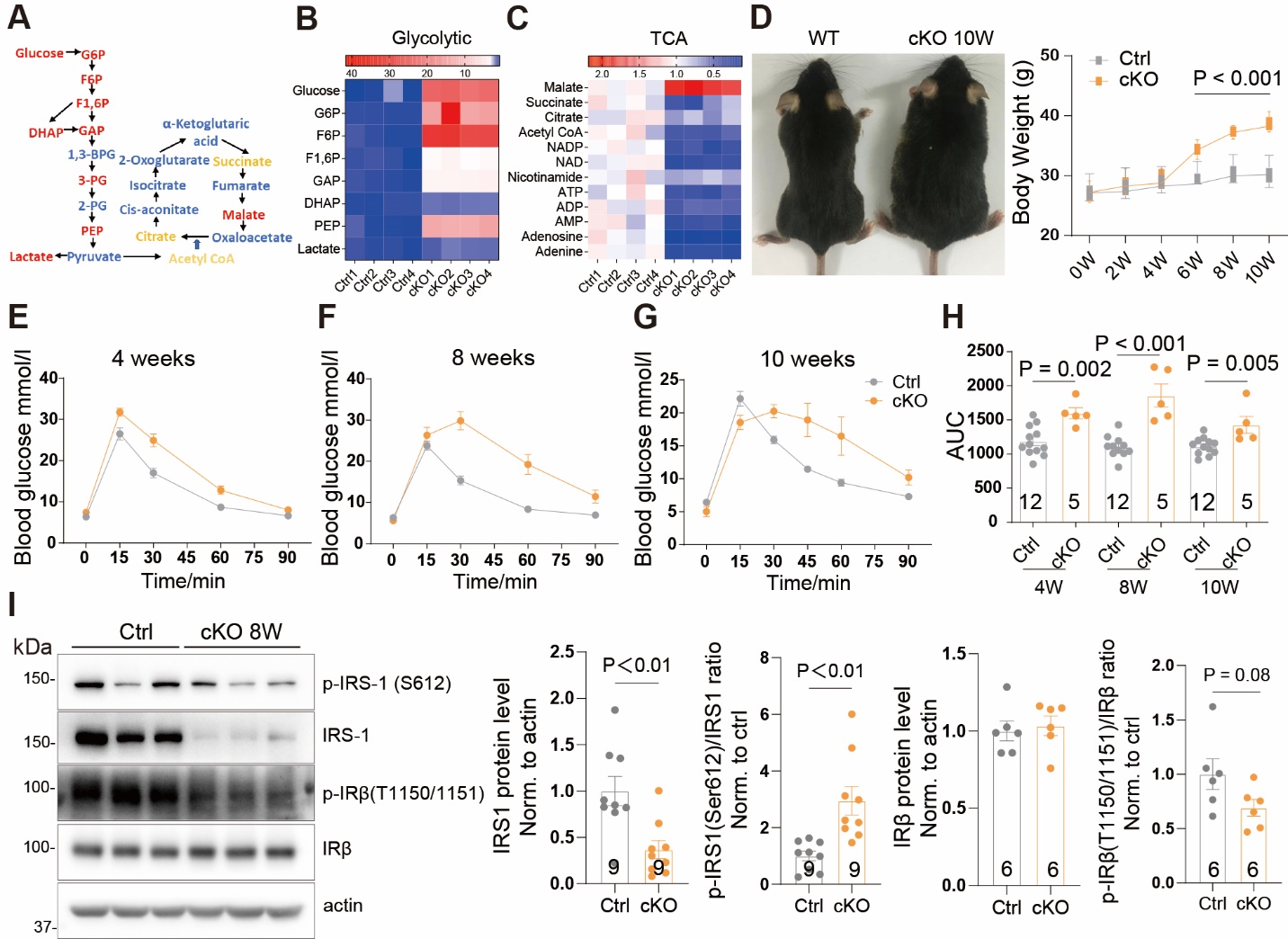

**Figure S7. The *Tpk*-cKO Mice manifest impairment in the homeostasis of cerebral and peripheral glucose metabolism.** Panel A shows schematics of glycolysis and oxidative phosphorylation pathways according to the non-targeting metabolomics studies. Red, yellow, and blue indicate increase, decrease, and no significant alteration, respectively, in the levels of examined components in cortical samples from the *Tpk*-cKO mice compared to control littermates in the 10^th^ week after tamoxifen treatment. Panel B shows heatmap of selected metabolic intermediates of glycolysis: glucose, glucose-6-phosphate (G6P), fructose-6-phosphate (F6P), fructose-1, 6-phosphate (F1, 6P), glyceraldehyde 3-phosphate (GAP), dihydroxy acetone phosphate (DHAP), lactate, and phosphoenolpyruvate (PEP). Panel C shows heatmap of selected metabolic intermediates of oxidative phosphorylation (tricarboxylic acid cycle, TCA) pathway: succinate, citrate, acetyl CoA, ATP, NAD, NADP, malate, and additional ones as indicated. Panel D shows representative images reflecting body weight in the 10^th^ week (*left*) and summary of body weights (*right*) for the control littermates and *Tpk*-cKO mice over the 10-week period after tamoxifen treatment. The *Tpk*-cKO mice (n = 6) showed progressive enhancement of body weight compared to their control littermates (n = 6), starting from the 6^th^ week. Panel E-G shows glucose tolerance tests of the control littermates and *Tpk*-cKO mice in the 4^th^ (E), 8^th^ (F), and 10^th^ (G) weeks after tamoxifen treatment. Panel H shows increased area under the curve (AUC, calculated from curves in E-G) of blood glucose levels in the *Tpk*-cKO mice compared to their control littermates. Panel I shows images of Western blots and quantification of IRS1, p-IRS1, IRβ, and p-IRβ protein levels in cortical tissues of the control littermates and *Tpk*-cKO mice in the 8^th^ week after tamoxifen treatment. Statistical analyses were performed using unpaired two-tailed Student’s t-tests. Summary data represent means ± SEM. The number of mice per group is shown in the statistical graph. P values are indicated above the bars or > 0.05 if not labeled.

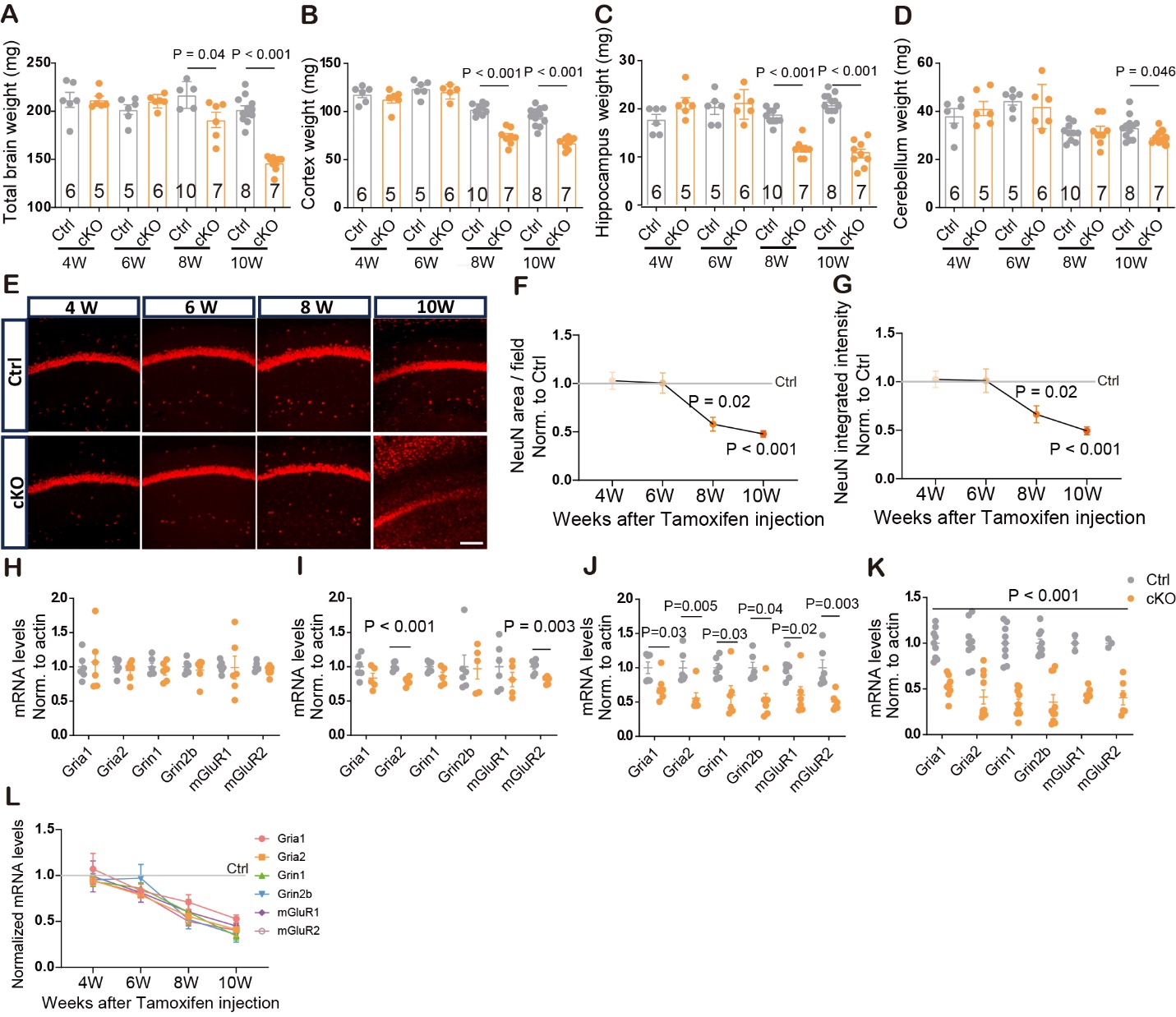

**Figure S8. Brain atrophy and progressive loss of synapses and neurons in the *Tpk*-cKO mice.** Panels A-D show quantifications of the weights of total brain (Panel A), cerebral cortex (Panel B), hippocampus (Panel C), and cerebellum (Panel D) of control littermates (Ctrl) and the cKO mice in the 4^th^, 6^th^, 8^th^, and 10^th^ weeks after tamoxifen treatment. There are no significant differences in the weights of total brain, cerebral cortex, hippocampus, and cerebellum between the two genotypes in the 4^th^ and 6^th^ weeks after tamoxifen treatment (P > 0.05 for all). There are significant decreases in weights of total brain, cerebral cortex, and hippocampus in the cKO mice compared to that in their control littermates in the 8^th^ and 10^th^ weeks after tamoxifen treatment. The weight of cerebellum in the cKO mice is slightly decreased compared to that in their control littermates only in the 10^th^ week (P = 0.046). Panels E-G show representative images (E) and quantifications of the areas (F) and integrated intensities (G) of NeuN-positive cells in hippocampal regions. Significant decreases were found in the *Tpk*-cKO mice compared to control littermates in the 8^th^ and 10^th^ weeks, but not in the 4^th^ and 6^th^ weeks. Panels H-K show mRNA levels of genes for synaptic glutamate receptors, *Gria*1, *Gria*2, *Grin*1, *Grin*2b, *mGluR*1, and *mGluR*2, in cortical samples of the *Tpk*-cKO mice were significantly decreased compared to that in their control littermates in the 6^th^ (I, n = 6, 5 for Ctrl and cKO, respectively, and the same convention herein), 8^th^ (J, n = 6, 6), and 10^th^ (K, n = 9, 10) weeks, but not in the 4^th^ week (H, n = 6, 6) after tamoxifen treatment. Panels L shows change tendency of mRNA levels for synaptic glutamate receptor genes plotted using values in (H-K). Summary data represent means ± SEM. Scale bars, 100 µm. The number of mice per group (n) is shown in the bar. P values are indicated above the bars or next to the data points (in panels f and g), or > 0.05 if not labeled (expect for panel L).

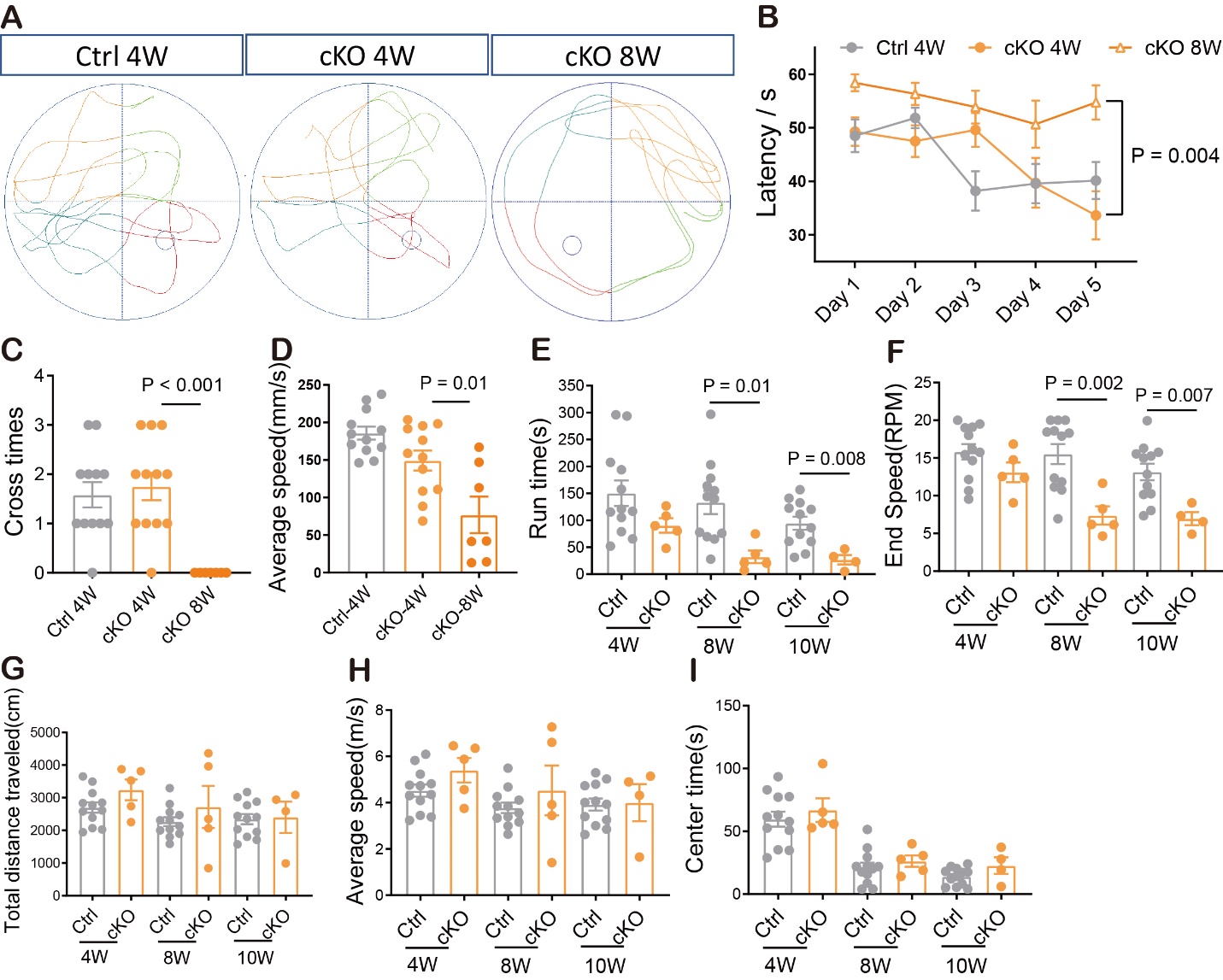

**Figure S9. The *Tpk-*cKO mice exhibit cognitive and motor dysfunctions.** Panel A shows representative plots depicting the paths of control littermates (Ctrl) and the *Tpk*-cKO mice swam to find the platform in the Morris water maze test in the 4^th^ (4W) and 8^th^ (8W) weeks after tamoxifen treatment. Panel B shows quantification of latency to mount the platform in the Morris water maze across the 5-day training period of control littermates (4W, n = 12) and the *Tpk*-cKO mice (4W, n = 11; 8W, n = 7). There is no significant difference in the latency between the genotypes in the 4^th^ week after tamoxifen treatment. A significant increase of the latency emerged in the *Tpk*-cKO mice in the 8^th^ week compared to the 4^th^ week after tamoxifen treatment (P = 0.004). Panels C and D show quantifications of the times of crossing the target site (Panel C) and average velocity in the water maze (Panel D) after retrieval of the platform on the testing day. While there are no significant differences between the two genotypes in the 4^th^ week, there are significant decreases in the *Tpk*-cKO mice in the 8^th^ week compared to that in the 4^th^ week after tamoxifen treatment. Panels E and F show results on run time (Panel E) and end speed (Panel F) of the rotarod test. While there are no significant changes in the *Tpk*-cKO mice (n = 5) compared to their control littermates (n = 12 for all weeks) in the 4^th^ week after tamoxifen treatment (P > 0.05), there are significant decreases in run time and end speed between the two genotypes in the 8^th^ and 10^th^ weeks after tamoxifen treatment. Panels G-I show results in total distance traveled (Panel G), average travel speed (Panel H), and time in the center (Panel I) of the open field test. No significant changes are found in the *Tpk*-cKO mice (n = 5) compared to their control littermates (n = 12, P > 0.05) in the 4^th^, 8^th^, and 10^th^ weeks after tamoxifen treatment. Summary data represent means ± SEM. P values are indicated above the bars or > 0.05 if not labeled.

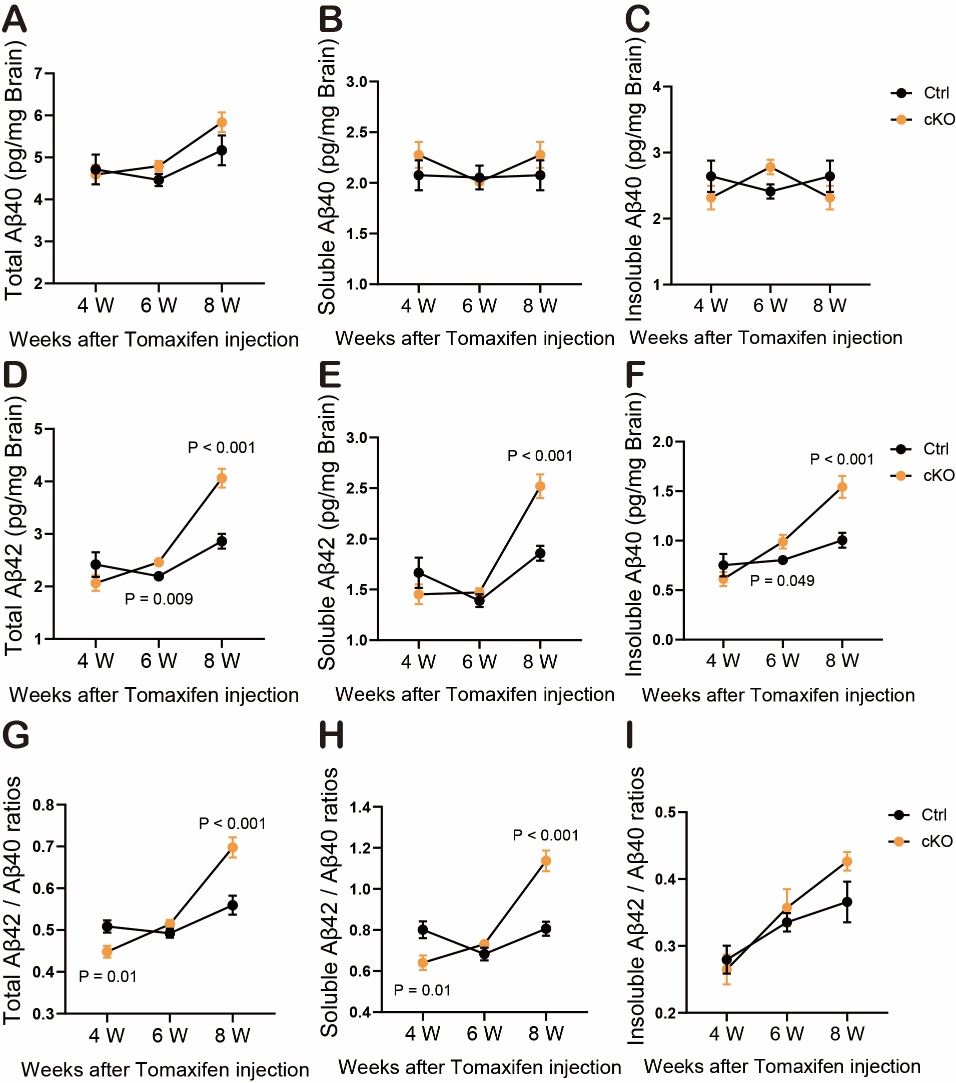

**Figure S10. Brain Aβ40 and Aβ42 levels detected by ELISA in control and the *Tpk*-cKO mice at 4^th^, 6^th^, and 8^th^ weeks after tamoxifen treatment.** Panels A-C show the levels of total (Panel A), soluble (Panel B), and insoluble (Panel C) Aβ40 in cortical samples from control littermates (Ctrl) and the *Tpk*-cKO mice in the 4^th^ (n = 6, 6, for Ctrl and cKO, respectively, and the same convention herein), 6^th^ (n = 6, 4), and 8^th^ (n = 6, 6) weeks after tamoxifen treatment. Panels D-F show the levels of total (Panel D), soluble (Panel E), and insoluble (Panel F) Aβ42 in cortical samples from control littermates and the *Tpk*-cKO mice in the 4^th^ (n = 6, 6), 6^th^ (n = 6, 4), and 8^th^ (n = 6, 6) weeks after tamoxifen treatment. Panels G-I show the Aβ42 / Aβ40 ratios in the total (Panel G), soluble (Panel H), and insoluble (Panel I) fractions of the cortical samples from control littermates (Ctrl) and the *Tpk*-cKO mice in the 4^th^ (n = 6, 6), 6^th^ (n = 6, 4), and 8^th^ (n = 6, 6) weeks after tamoxifen treatment. Summary data represent means ± SEM. P values are indicated or > 0.05 if not labeled.

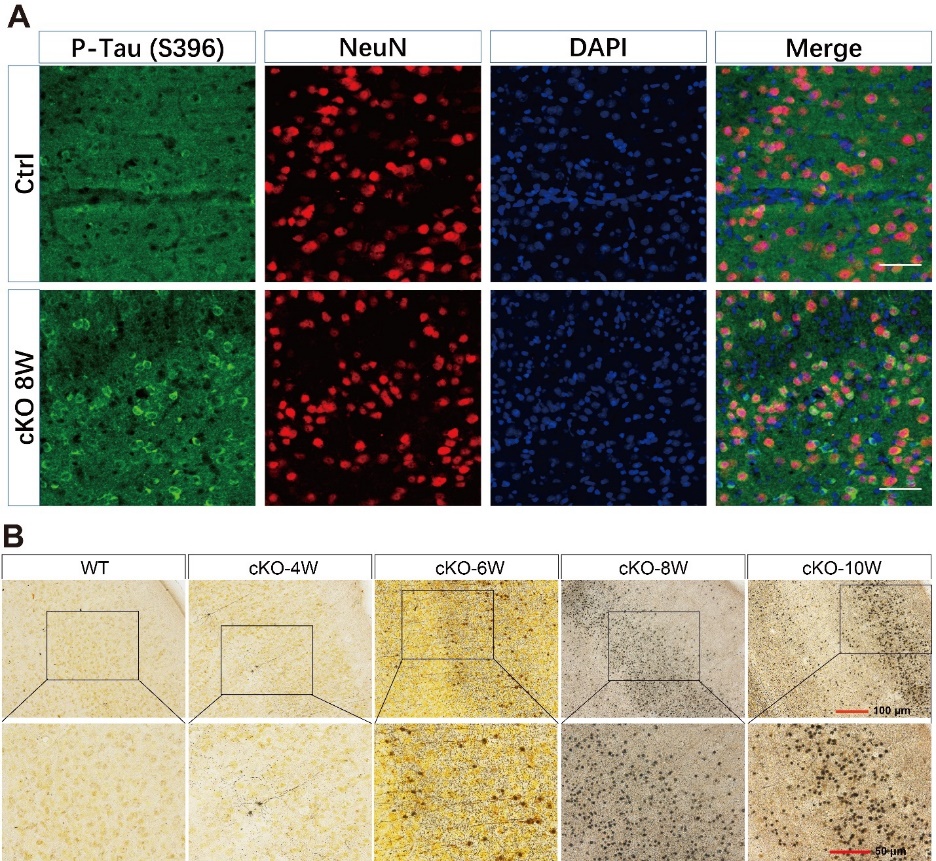

**Figure S11. Neuronal Tau hyperphosphorylation and neurofibrillary tangles in the *Tpk*-cKO mice.** Panel A shows representative images of phosphorylated Tau stained by the P-Tau (S396) antibody, which co-existed with neurons stained by the NeuN antibody in cortical samples of the *Tpk*-cKO mice in the 8^th^ week after tamoxifen treatment. Panel B shows representative images of Gallyas silver staining of cortical samples from control littermates and the *Tpk*-cKO mice in the 4^th^, 6^th^, 8^th^, and 10^th^ weeks after tamoxifen treatment. Scale bars, 100 µm in the upper row and 50 µm in the lower row.

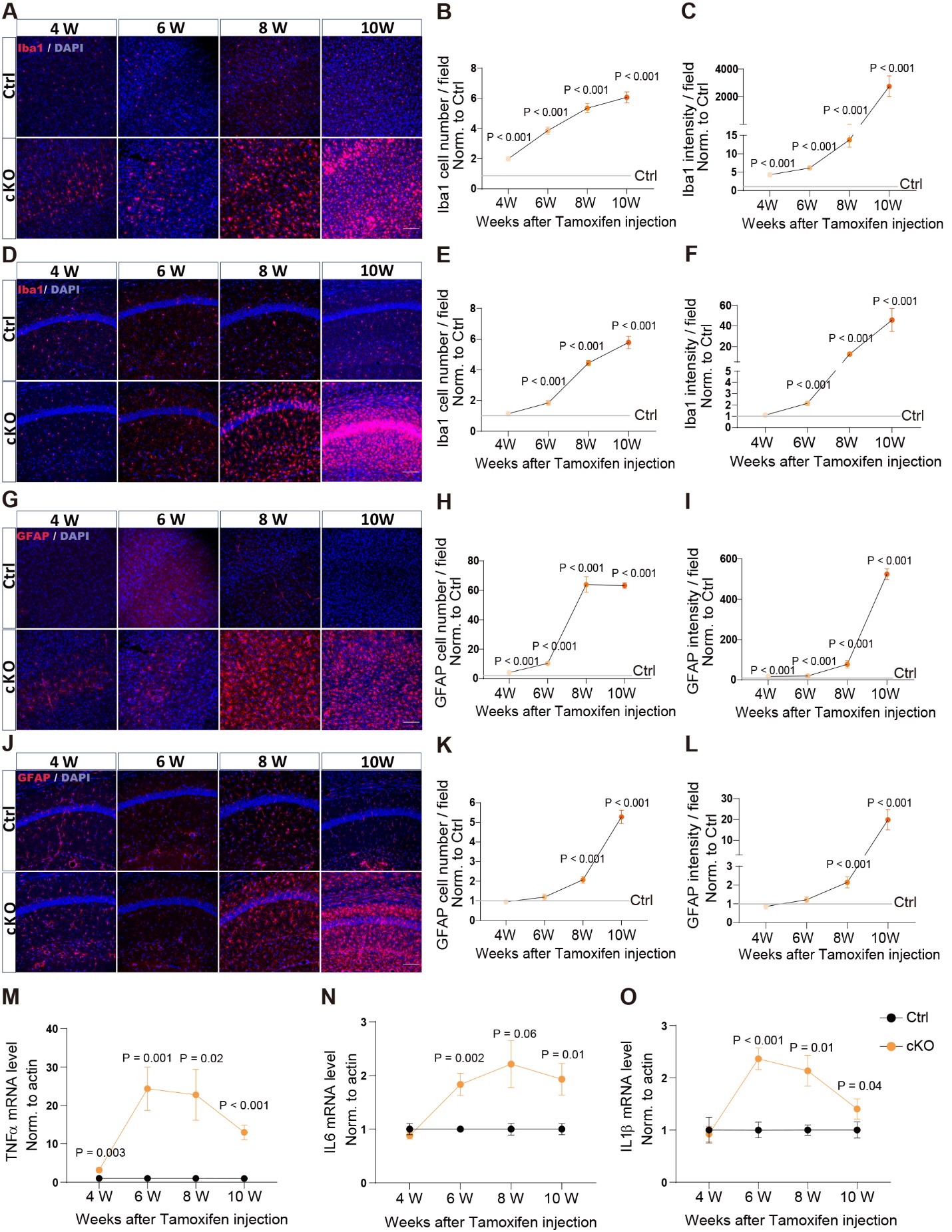

**Figure S12. Glial activation and alterations of inflammatory cytokines in brains of the *Tpk*-cKO mice.** Panels A-C show cortical microglia in control littermates and the cKO mice in the 4^th^ (n = 6, 6), 6^th^ (n = 6, 5), 8^th^ (n = 5, 7), and, 10^th^ (n = 5, 4) weeks after tamoxifen treatment. Representative images (Panel A) and quantifications on the numbers (Panel B) and intensities (Panel C) of Iba1-positive cells are shown. Panels D-F show hippocampal microglia in control littermates and the *Tpk*-cKO mice in the 4^th^ (n = 6, 6), 6^th^ (n = 6, 5), 8^th^ (n = 5, 7), and 10^th^ (n = 5, 4) weeks after tamoxifen treatment. Representative images (Panel D) and quantifications on the numbers (Panel E) and intensities (Panel F) of Iba1-positive cells are shown. Panels G-I show cortical astrocytes in control littermates and the *Tpk*-cKO mice in the 4^th^ (n = 6, 6), 6^th^ (n = 7, 5), 8^th^ (n = 5, 7), and 10^th^ (n = 5, 4) weeks after tamoxifen treatment. Representative images (Panel G) and quantifications on the numbers (Panel H) and intensities (Panel I) of GFAP-positive cells are shown. Panels J-L show hippocampal astrocytes in control littermates and the *Tpk*-cKO mice in the 4^th^ (n = 6, 6), 6^th^ (n = 6, 5), 8^th^ (n = 5, 7), and 10^th^ (n = 5, 4) weeks after tamoxifen treatment. Representative images (Panel J) and quantifications on the numbers (Panel K) and intensities (Panel L) of GFAP-positive cells are shown. Panels M-O show the mRNA levels of inflammatory cytokines: TNFα (Panel M), IL6 (Panel N), and IL1β (Panel O) measured by RT-qPCR in brain samples of control littermates and the *Tpk*-cKO mice in the 4^th^ (n = 6, 6), 6^th^ (n = 6, 5), 8^th^ (n = 6, 7), and 10^th^ (n = 9, 10) weeks after tamoxifen treatment. Summary data represent means ± SEM. Scale bars, 100 µm. P values are indicated above the data points or > 0.05 if not labeled.

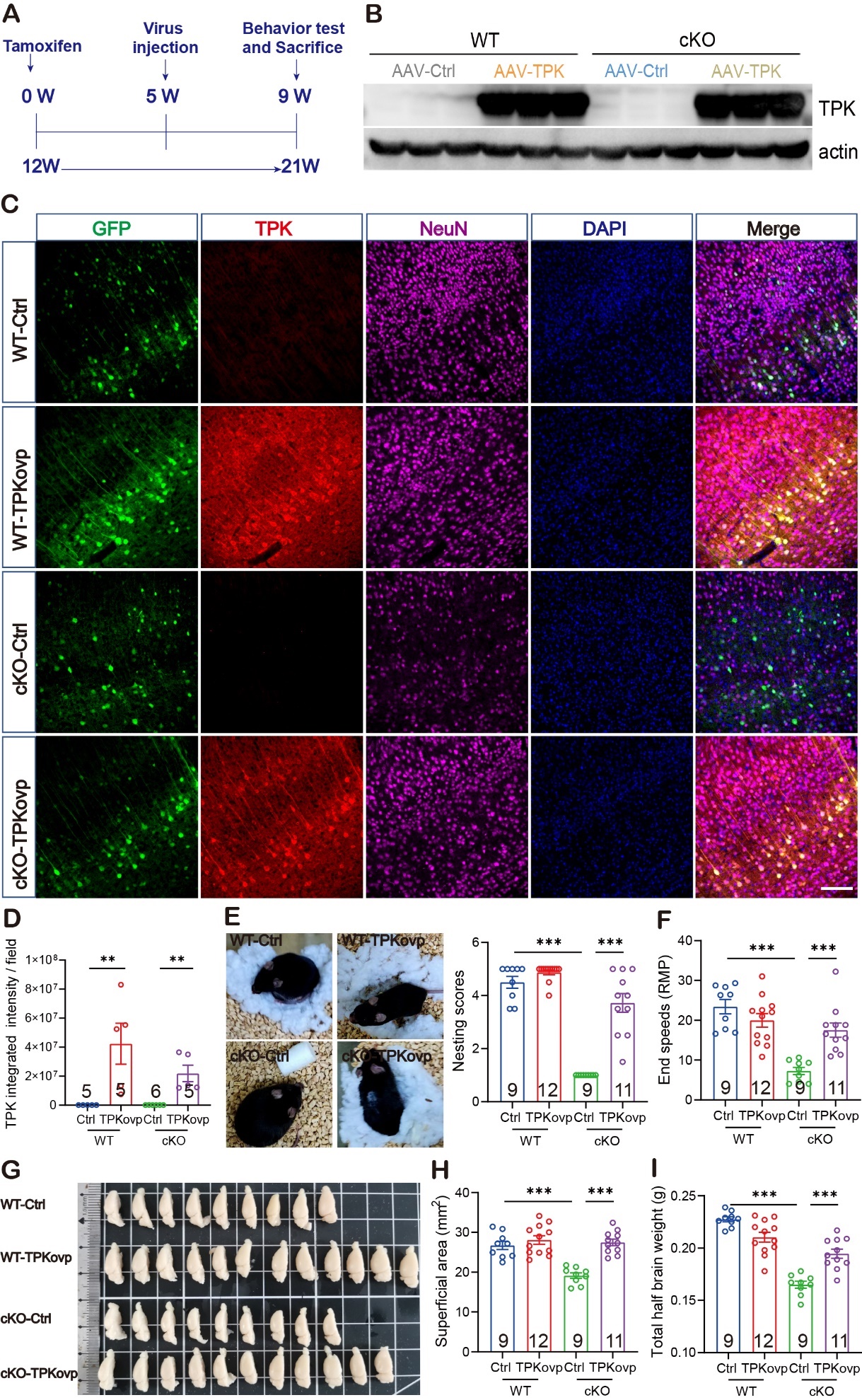

**Figure S13. Alleviation of cognitive dysfunction and brain atrophy of the *Tpk*-cKO mice through brain delivery of *Tpk*.** Panel A shows AAV-CAP-B10 viruses expressing GFP or TPK were intravenously injected to the *Tpk*-cKO mice in the 5^th^ week, and behavioral and other tests were performed in the 9^th^ week after tamoxifen treatment. Panel B shows images of Western blots of TPK protein in cortical samples. Panels C and D show representative images and quantification of cortical TPK protein detected by immunofluorescent staining, showing successful expression of TPK in brain neurons of the *Tpk*-cKO and wild-type (WT) mice. Panel E shows Nest-building assessed after 48 hours of housing showing impairment in the *Tpk*-cKO mice in the 9^th^ week after tamoxifen treatment compared to control littermates, which was rescued by restoration of TPK in the brain. Panel F shows in rotarod test, end speed of running was significantly decreased in the *Tpk*-cKO mice, which was rescued by restoration of TPK in the brain. Panels G and H show the decrease of brain superficial areas in the *Tpk*-cKO mice was rescued by restoration of TPK in the brain. Panel I shows the decrease in total brain weight was rescued by restoration of TPK in the brain. Summary data represent means ± SEM. Scale bars, 100 µm. P values are indicated above the bars for *P < 0.05, **P < 0.01, ***P < 0.001 or > 0.05 if not labeled.

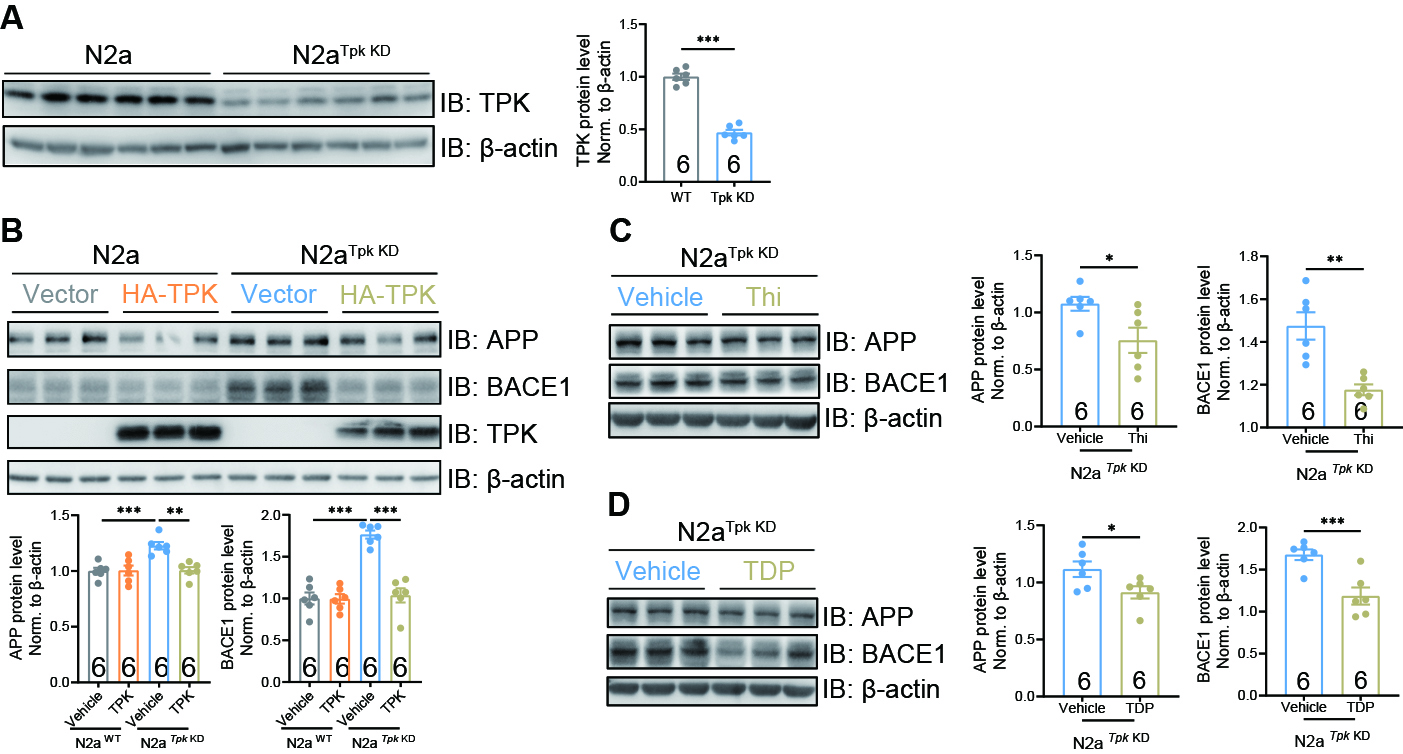

**Figure S14. *Tpk* knockdown increases APP and BACE1 expression in N2a cells, which is rescued by TPK overexpression or supplementation of TDP or thiamine (Thi).** Panel A shows images and quantification of TPK detected by Western blotting, showing that TPK RNAi significantly down-regulated the level of TPK protein in N2a cells (stable N2a*^Tpk^* ^KD^ cell line) compare to wide-type N2a (N2a^WT^) cells. Panel B shows representative images and quantification of APP and BACE1 proteins expressed in the N2a*^Tpk^* ^KD^ and N2a^WT^ cells detected by Western blotting, showing the rescue by TPK overexpression. Panel C shows representative images and quantification of APP and BACE1 proteins in the N2a*^Tpk^* ^KD^ cells by Western blotting, showing the rescue by thiamine treatment (0.1 mg/ ml, 48 hours). Panel D shows representative images and quantification of APP and BACE1 proteins in the N2a*^Tpk^* ^KD^ cells by Western blotting, showing the rescue by TDP treatment (0.1 mg / ml, 48 hours). Summary data represent means ± SEM. P values are indicated above the bars for *P < 0.05, **P < 0.01, ***P < 0.001 or > 0.05 if not labeled.

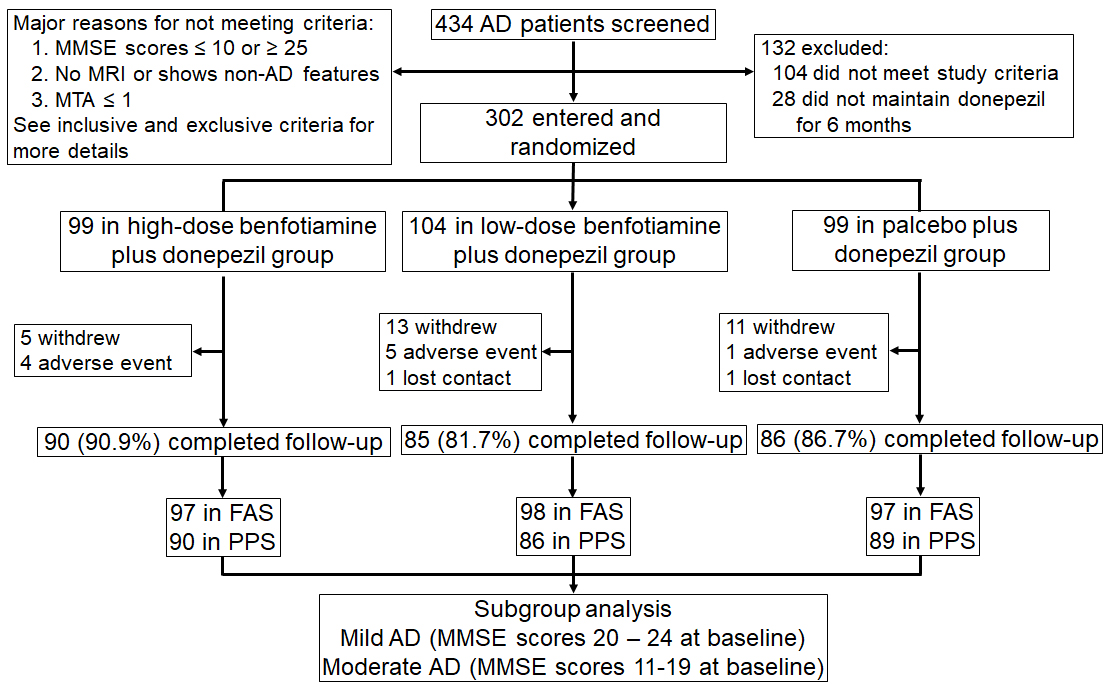

**Figure S15. Screening, randomization, and trial completion.** A total of 434 mild-to-moderate Alzheimer’s disease (AD) patients, as defined by the Mini-Mental State Examination (MMSE) scores of 11 to 24, were screened and 302 patients were randomly assigned into high-dose (n = 99), low-dose (n = 104), and placebo (n = 99) groups. FAS: Full analysis set; PPS: Per protocol set; MTA: medial temporal lobe atrophy score.

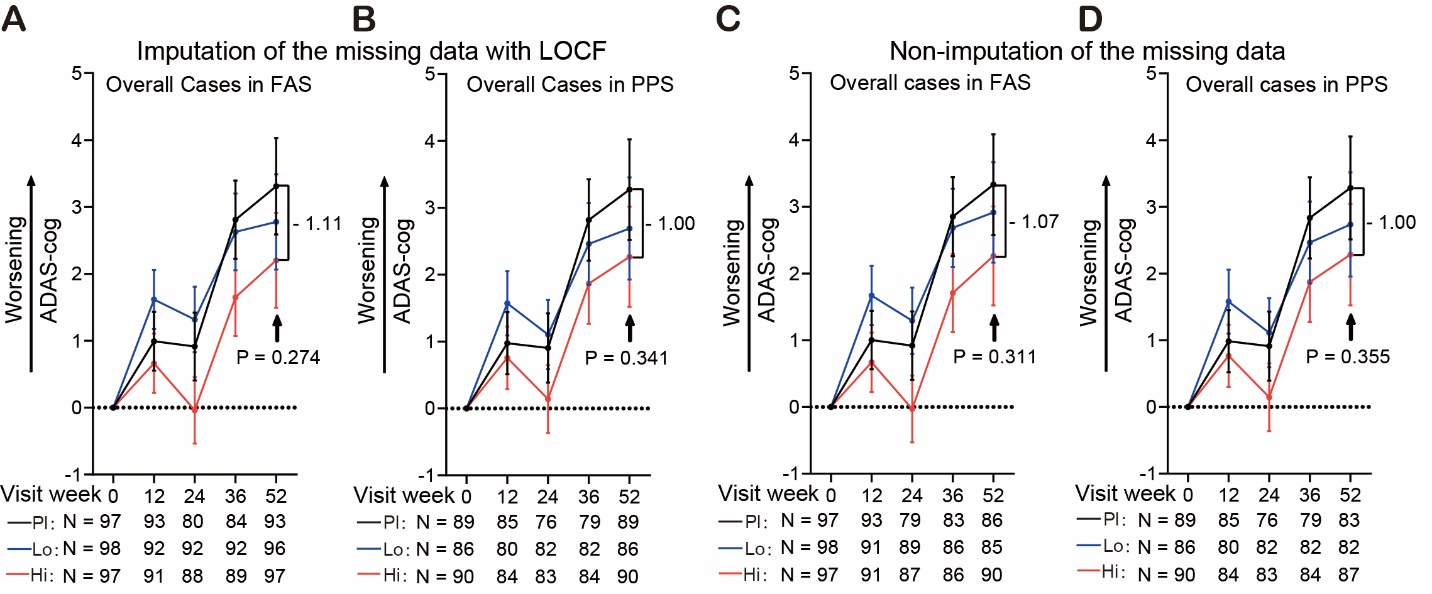

**Figure S16. Mean changes of ADAS-cog scores for all AD patients with LOCF or without imputation of the missing data.** Panels A and B show mean changes of ADAS-cog scores at the indicated time points from the baseline for all AD patients (overall cases) among high-dose benfotiamine (600 mg daily, Hi), low-dose benfotiamine (300 mg daily, Lo), and placebo (Pl) groups in full analysis set (FAS, A) and per protocol set (PPS, B). There was a dose-relationship but no statistical difference in the mean changes of the ADAS-cog scores among the three groups in both FAS and PPS by MMRM analyses. The missing data of the ADAS-cog score were imputed using the last observation carried forward (LOCF) method. Panels C and D show MMRM analysis of the same data as in (A) and (B) without imputation of the missing ADAS-cog scores. There was a dose-relationship but no statistical difference in the mean changes of the ADAS-cog scores among the three groups in both FAS and PPS.

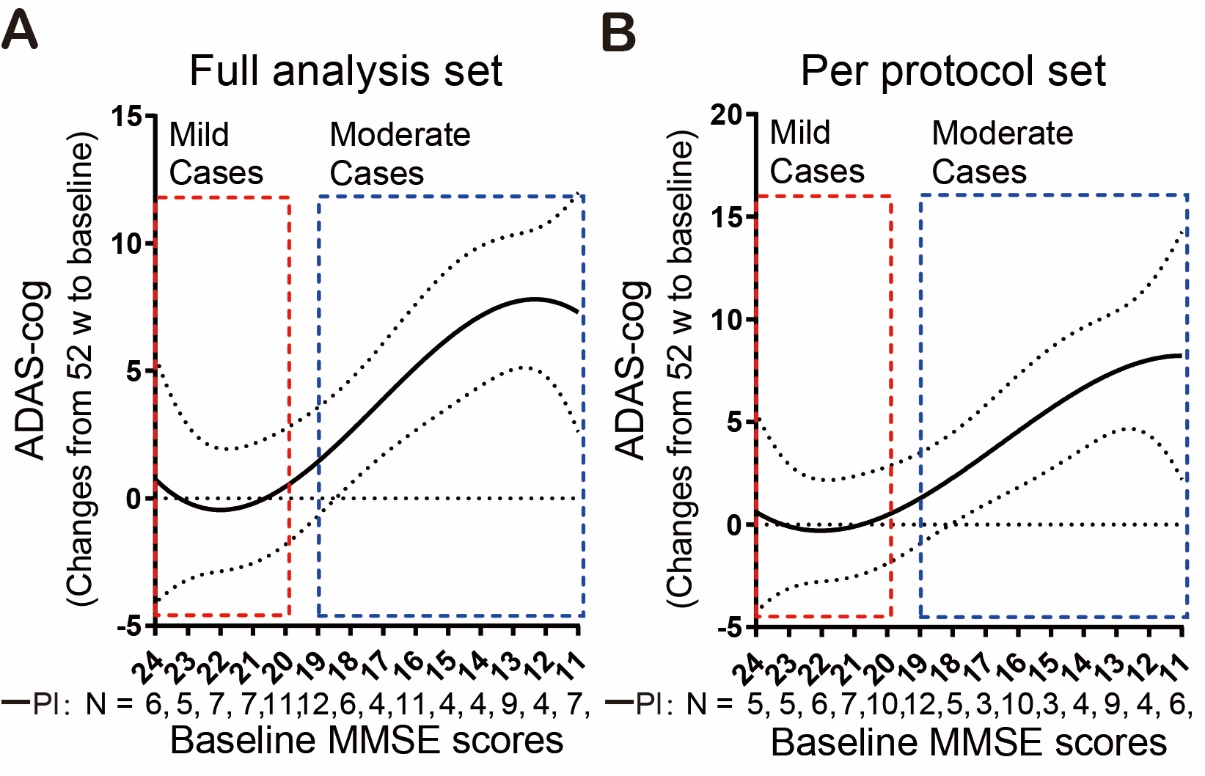

**Figure S17. Changes of ADAS-cog scores of AD patients at week 52 from baseline in the placebo group in FAS (A) and PPS (B), plotted against their MMSE scores at the baseline.**

**Table S1. Expression of genes associated with thiamine metabolism in brain samples of patients with AD and other neurodegenerative disorders.**

| GEO  Super-Series | GEO  Sub-Series | Disease | Samples | Controls | Cases | Thiamine metabolism genes | | | |
| --- | --- | --- | --- | --- | --- | --- | --- | --- | --- |
|  |  |  |  |  |  | ***TPK*** | ***SLC19A2*** | ***SLC19A3*** | ***SLC25A19*** |
| GSE95587 | **/** | AD | 117 | 33 | 84 | **↓** | **ns** | **ns** | **ns** |
| GSE15222 | **/** | AD | 241 | 135 | 106 | **↓** | **ns** | **ns** | **ns** |
| GSE44772 | GSE44768 | AD | 230 | 101 | 129 | **↓** | **ns** | **↑** | **↑** |
|  | GSE44770 |  | 230 | 101 | 129 | **↓** | **↑** | **↑** | **↑** |
|  | GSE44771 |  | 230 | 101 | 129 | **↓** | **ns** | **↑** | **ns** |
| GSE13162 | **/** | FTLD | 56 | 17 | 39 | **ns** | **ns** | **ns** | **ns** |
| GSE20295 | GSE20168 | PD | 29 | 15 | 14 | **ns** | **ns** | **ns** | **ns** |
|  | GSE20291 |  | 35 | 20 | 15 | **ns** | **ns** | **ns** | **ns** |
|  | GSE20292 |  | 29 | 18 | 11 | **ns** | **ns** | **ns** | **ns** |
| GSE3790 | GSE3790A | HD | 201 | 87 | 114 | **ns** | **ns** | **ns** | **ns** |
|  | GSE3790B |  | 203 | 89 | 114 | **ns** | **ns** | **ns** | **ns** |
| GSE18920 | **/** | ALS | 22 | 10 | 12 | **ns** | **ns** | **ns** | **ns** |

(**↓**indicates significant reduction with P < 0.01, **↑**indicates significant increase with P < 0.05, ns indicates no significant change).

**Table S2. Characteristics of the AD patients and control subjects for TPK protein analysis.**

| **Subjects** | **Ethnicity** | **Gender** | **Death cause** | **Age (years)** | **Brain Area** |
| --- | --- | --- | --- | --- | --- |
| Controls |  |  |  |  |  |
| 1743 | Caucasian | Female | ASCVD | 44 | FC |
| 794 | Caucasian | Female | ASCVD | 51 | FC |
| 4789 | Caucasian | Female | accident, exsanguination | 72 | FC |
| 1569 | Caucasian | Female | drowning complicating HASCVD | 77 | FC |
| 4546 | Caucasian | Female | HASCVD | 86 | FC |
| 1170 | Caucasian | Male | cardiac arrhythmia/endocarditis | 58 | FC |
| 4263 | Caucasian | Male | cardiac arrest | 61 | FC |
| 1213 | Caucasian | Male | ASCVD | 67 | FC |
| 4735 | Caucasian | Male | COPD | 73 | FC |
| Alzheimer's Disease |  |  |  |  |  |
| 4833 | Caucasian | Female | bronchopneumonia (hypoxia) | 59 | FC |
| 1720 | Caucasian | Female | complications of disorder | 63 | FC |
| 1212 | Caucasian | Female | complications of disorder | 72 | FC |
| 1463 | Caucasian | Female | complications of disorder | 90 | FC |
| 4556 | Caucasian | Female | complications of disorder | 70 | FC |
| 4759 | Caucasian | Female | complications of disorder | 77 | FC |
| 4854 | Caucasian | Male | complications of disorder | 54 | FC |
| 1946 | Caucasian | Male | coronary artery disease | 68 | FC |
| 1625 | Caucasian | Male | complications of disorder | 70 | FC |
| 1780 | Caucasian | Male | complications of disorder | 72 | FC |
| 4512 | Caucasian | Male | complications of disorder | 77 | FC |
| 1252 | Caucasian | Male | complications of disorder | 78 | FC |
| Abbreviations: FC, Frontal Cortex; ASCVD, Atherosclerotic Cardiovascular Disease; HASCVD, Hypertensive Arteriosclerotic Cardiovascular Disease; COPD, Chronic Obstructive Pulmonary Disease | | | | | |

| Table S3. Baseline Characteristics of the Participants (FAS Population) | | | | |
| --- | --- | --- | --- | --- |
|  | **High-dose Benfotiamine**  **plus Donepezil**  **(n = 97)** | **Low-dose Benfotiamine**  **plus Donepezil**  **(n = 98)** | **Placebo plus**  **Donepezil**  **(n = 97)** | **Total**  **(n = 292)** |
| Age — yr (SD) | 66.97 (6.70) | 67.50 (7.84) | 67.76 (7.70) | 67.41 (7.41) |
| Sex — no. (%) | | | | |
| Male | 39 (40.21) | 37 (37.76) | 39 (40.21) | 115 (39.38) |
| Female | 58 (59.79) | 61 (62.24) | 58 (59.79) | 177 (60.62) |
| BMI (kg/m2) | 22.49 (3.17) | 23.01 (3.18) | 22.45 (3.12) | 22.65 (3.16) |
| Race — no. (%) | | | | |
| Han | 95 (97.94) | 97 (98.98) | 97 (100.00) | 289 (98.97) |
| Others | 2 (2.06) | 1 (1.02) | 0 (0.00) | 3 (1.03) |
| ^&^APOE ε4 carrier—no.(%) | 48 (49.48) | 45 (45.92) | 46 (47.42) | 139 (47.60) |
| Non-carriers—no. (%) | 44 (45.37) | 44 (44.90) | 43 (44.33) | 131 (44.86) |
| Unknown—no. (%) | 5 (5.15) | 9 (9.18) | 8 (8.25) | 22 (7.54) |
| Education —no. (%) | | | | |
| Primary school | 21 (21.65) | 18 (18.37) | 17 (17.53) | 56 (19.18) |
| Secondary school | 55 (56.70) | 48 (48.98) | 55 (56.70) | 158 (54.11) |
| College & above | 21 (21.65) | 32 (32.65) | 25 (25.77) | 78 (26.71) |
| MMSE scores (SD) | 17.88 (3.56) | 17.51 (4.04) | 17.71 (3.85) |  |
| ^$^ Moderate case number- no. (%)  (MMSE 11 ~19 scores) | 64 (65.98) | **^$^** 58 (59.18) | 61 (62.89) | 183 (62.67) |
| Mild case number- no. (%)  (MMSE 20 ~24 scores) | 33 (34.02) | 37 (37.76) | 36 (37.11) | 106 (37.33) |
| ADAS-cog (SD) | 22.4 (7.93) | 23.2 (9.60) | 22.1 (8.51) |  |
| ADCS-ADL (SD) | 63.54 (6.82) | 61.96 (9.57) | 62.73 (9.44) |  |
| CDR n (%) |  |  |  |  |
| CDR (0.5) | 49 (50.52) | 37 (37.76) | 43 (44.33) | 129 (44.18) |
| CDR (1) | 47 (48.45) | 57 (58.16) | 50 (51.55) | 154 (52.74) |
| CDR (2) | 1 (1.03) | 4 (4.08) | 4 (4.12) | 9 (3.08) |
| NPI-NH (SD) | 6.51 (8.38) | 7.09 (8.86) | 6.07 (7.37) |  |
| NPI-D (SD) | 2.92 (4.09) | 2.59 (3.61) | 2.78 (4.19) |  |

Data are no. (%) or mean (SD). FAS: the full-analysis set; BMI: Body-mass index; APOE: Apolipoprotein E; MMSE: Mini-Mental State Examination; ADAS-cog: the 11-item cognitive subscale of the Alzheimer’s Disease Assessment Scale; ADCS-ADL: Alzheimer’s Disease Cooperative Study-activities of daily living; CDR: Clinical Dementia Rating; NPI–NH: Neuropsychiatric Inventory–Nursing Home version; NPI-D: NPI caregiver distress scale.

**&**: Ninety-two participants in high-dose benfotiamine plus donepezil group, 89 in low-dose benfotiamine plus donepezil group, and 89 in placebo plus donepezil received APOE genotype measurement.

**$**: Three cases in low-dose benfotiamine plus donepezil group were not included in the subgroup analyses because their MMSE scores deteriorated to 10 at baseline.
