## Supplementary material for "Thiamine pyrophosphokinase-1 deficiency in neurons drives Alzheimer’s multiple pathophysiological alterations": Methods

**Materials and Methods**

**GEO datasets download**

To detect the expression changes of all four known genes associated with thiamine metabolism and TDP, including *TPK*, *SLC19A2*, *SLC19A3*, and *SLC25A19*, a total of 7 public RNA-seq and microarray datasets for brain samples of patients with AD, FTLD, PD, HD, and ALS from the GEO database were analyzed. The datasets are GSE95587, GSE15222, GSE44772 (44768, 44770, 44771), GSE13162, GSE20295 (20168, 20291, 20292), GSE3790 (3790A, 3790B), and GSE18920. R package limma was used to perform differential expression (DE) analysis for all genes in each GEO dataset, and P values were adjusted using Benjamini-Hochberg method. FDR cut-off of 0.05 was used to select significantly differentially expressed genes of interest.

GSE95587 is an Illumina HiSeq 2500 RNA-seq dataset, measuring fusiform gyrus tissue samples of 117 subjects, including 84 AD patients and 33 neurologically normal age-matched control subjects with RNA integrity scores of at least 5 and post-mortem intervals no greater than 5 hours. The detailed data can be found in the paper published in Cell Reports ^1^. This dataset was downloaded from https://www.ncbi.nlm.nih.gov/ geo/download/?acc=GSE95587&format=file.

GSE44772 ^2^ is a Rosetta/Merck Human 44k 1.1 microarray dataset, which is composed of the following Sub-Series: GSE44768, GSE44770 and GSE44771.

GSE44768 measures cerebellar tissue samples of 230 subjects, including 129 late onset AD (LOAD) patients and 101 non-demented healthy controls. This dataset was downloaded from https://ftp.ncbi.nlm.nih.gov/geo/series/GSE44nnn/GSE44768/ matrix.

GSE44770 measures tissue samples of dorsolateral prefrontal cortex in the same 230 subjects. This dataset was downloaded from https://ftp.ncbi.nlm.nih.gov/geo/series/ GSE44nnn/GSE44770/matrix.

GSE44771 measures tissue samples of visual cortex in the same 230 subjects. This dataset was downloaded from https://ftp.ncbi.nlm.nih.gov/geo/series/GSE44nnn/ GSE44771/matrix.

GSE15222 ^3^ is a Sentrix Human Expression BeadChip dataset, measuring postmortem brain samples of 363 subjects, including 176 LOAD cases and 187 neuropathologically normal controls. Due to that the datasets were mixed with data from multiple brain regions, only the temporal cortex dataset with 135 neuropathologically normal controls and 106 AD cases was used for further analysis (The detailed sample information is shown in the table below, cited from references). This dataset was downloaded from https://ftp.ncbi.nlm.nih.gov/geo/series/GSE15nnn/GSE15222/ matrix.

Sample description (from the reference entitled Integrated systems approach identifies genetic nodes and networks in late-onset Alzheimer's disease) ^2^.

|  | Control cohort | Case cohort |
| --- | --- | --- |
| % female | 45% | 50% |
| average age | 81 | 84 |
| age range | 65-100 | 68-102 |
| % frontal cortex | 21% | 18% |
| % temporal cortex | 73% | 60% |
| % parietal cortex | 2% | 10% |
| % cerebellar cortex | 3% | 13% |
| of different hybridization dates | 9 | 9 |
| average # of samples per hybridization | 21 | 19 |
| number of institute sources | 18 | 16 |
| average number of samples per institute | 11 | 11 |
| average PMI | 10 hrs | 9 hrs |
| average transcriptome detection rate per sample | 77% | 76% |

GSE13162 ^4^ is an Affymetrix Human Genome U133A 2.0 Array dataset, measuring postmortem frontal samples of 56 subjects, including 39 FTLD patients and 17 normal controls. This dataset was downloaded from https://www.ncbi.nlm.nih.gov/geo/ download/?acc=GSE13162&format=file.

GSE20295 ^5^ is an Affymetrix Human Genome U133A Array dataset, which is composed of the following Sub-Series: GSE20168, GSE20291 and GSE20292.

GSE20168 measures prefrontal tissue samples of 29 subjects, including 14 PD patients and 15 normal controls. This dataset was downloaded from https://www.ncbi.nlm.nih.gov/geo/download/?acc=GSE20168&format=file.

GSE20291 measures putamen tissue samples of 35 subjects, including 15 PD patients and 20 normal controls. This dataset was downloaded from https://www.ncbi. nlm.nih.gov/geo/ download/?acc=GSE20291&format=file.

GSE20292 measures substantia nigra tissue samples of 29 subjects, including 11 PD patients and 18 normal controls. This dataset was downloaded from https://www.ncbi. nlm.nih.gov/geo/ download/?acc=GSE20292&format=file.

GSE3790 ^6^ is an Affymetrix Human Genome Array dataset. This dataset was downloaded from https://www.ncbi.nlm.nih.gov/geo/download/?acc=GSE3790& format=file. Samples of this dataset were profiled using two different Array platforms (Affymetrix Human Genome U133A Array and Affymetrix Human Genome U133B Array). Therefore, samples were separately analyzed in different platforms. The dataset from Affymetrix Human Genome U133A Array was named GSE3790A, and that from Affymetrix Human Genome U133B Array named GSE3790B. GSE3790A measures postmortem frontal cortex, cerebellum, and caudate nucleus samples of 201 subjects, including 114 HD patients and 87 normal controls. GSE3790B measures postmortem frontal cortex, cerebellum, and caudate nucleus samples of 203 subjects, including 114 HD patients and 89 normal control subjects.

GSE18920 ^7^ measures 22 total lumbar spinal cord samples including 12 sporadic ALS and 10 control subjects. Because no annotation files could be identified for this dataset, we obtained expression data for *TPK*, *SLC19A2*, *SLC19A3*, and *SLC25A19* from the well analyzed datasets (http://research-pub.gene.com/ BrainMyeloidLandscape/) ^1^.

**GEO data processing**

Affymetrix microarray data GSE13162, GSE20168, GSE20291, GSE20292, GSE3790A, and GSE3790B were processed (including background correction, normalization, and expression quantification) using the Robust Multi-Array Average (RMA) method implemented in R package *affy*1 (version 1.48.0). For GSE13162, GSE20168, GSE20291, GSE20292, and GSE3790A, gene annotation files were downloaded from https://www.ncbi.nlm.nih.gov/geo/query/acc.cgi?acc=GPL96, and for GSE3790B, the gene annotation file was downloaded from https://www.ncbi.nlm.nih.gov/geo/query/acc.cgi?acc=GPL97. Expression levels of multiple probe sets corresponding to the same gene were averaged to represent the expression level of the gene.

**Illumina HiSeq 2500 data processing**

Data of GSE95587 have already been processed using "nRPKM" normalization. This is similar to RPKM normalization but includes an extra step for size factor adjustment as previously described ^1^.

**Sentrix Human Expression BeadChip data processing**

We used the processed data with probe set expression levels of GEO15222. Missing values were inputted with the mean value across all samples using locally developed Perl code. The gene annotation file was downloaded from https://www.ncbi.nlm.nih.gov/geo/query/acc.cgi?acc=GPL2700. Expression levels of multiple probe sets corresponding to the same gene were averaged to represent the expression level of the gene. Genes with negative expression values were discarded.

**Rosetta/Merck Human 44k 1.1 microarray data processing**

Gene expression profiling was downloaded from GEO44768, GEO44770, and GEO44771. Gene expression was reported as the mean-log ratio of individual microarray intensities relative to average intensities of all samples. Detailed data processing steps are described in the Experimental Procedures of the reference ^1^.

**Human brain tissues and western blot analysis**

Cortical samples of AD patients and control subjects were obtained from the Department of Pathology, Columbia University (Table S2). The average ages of 9 controls were 65.4 ± 4.4 and 12 AD patients were 70.8 ± 2.7. For Western blotting, frozen cortical tissues were homogenized using a polytron homogenizer in a RIPA buffer supplemented with complete mini protease inhibitor cocktail tablet (Roche, 11836170001). Proteins were resolved by SDS polyacrylamide gel electrophoresis (SDS-PAGE) on 10% tris-glycine gels and transferred to nitrocellulose membranes. Membranes were blocked in 5% skim milk at room temperature for 1 hour and sequentially blotted with the primary antibody for TPK (Abcam, ab249546, 1:1000), followed by IRDye fluorescence labelled goat anti-rabbit IgG secondary antibodies (LI-COR Biosciences, 926-68021, 1:5000). β-actin (Sigma, A5441, 1: 5000) was used as the loading control.

**Preparation of oligomeric Aβ and cell treatment**

Human Aβ42 peptides, treated with 1,1,1,3,3,3-hexafluoro-2-propanol (HFIP), were purchased from Chinapeptides Co., LTD. (Qyaobio). The monomer Aβ lyophilized powder was dissolved to 5 mM in anhydrous dimethyl sulfoxide (anhydrous DMSO; Sigma-Aldrich) and stored at -80℃. Oligomeric Aβ was prepared by diluting the 5 mM Aβ42 in DMSO with cold DMEM/F12 cell culture media to a final concentration of 100 μM, followed by vortex for 15s and incubation at 4°C for 24 hours. On the 10^th^ day, the mature neurons were treated with 3 μM and 10 μM oligomeric Aβ. N2a cells were treated with 10 μM and 30 μM oligomeric Aβ. After 48 hours of treatment, the neurons were collected and homogenized using a polytron homogenizer in a radioimmunoprecipitation assay (RIPA) buffer (1% Triton X-100, 1% sodium deoxycholate, 0.2% sodium dodecyl sulfate (SDS), 0.15 M NaCl, 0.05 M Tris-HCl, pH 7.2) supplemented with complete mini protease inhibitor cocktail tablet (Roche, 11836170001) for protein detection using Western blotting.

**Animals**

All animal care and experimental procedures were approved by the Medical Experimental Animal Administrative Committee of Fudan University and by the Institutional Animals Care and Use Committee of the Institute of Neuroscience, Chinese Academy of Sciences. The amyloid precursor protein/presenilin-1 (APP/PS1; strain 4462) double transgenic mice were purchased from The Jackson Laboratory (Bar Harbor, ME, USA) and used as heterozygotes.

**Intraperitoneal injection of streptozotocin in mice**

Streptozotocin (STZ; Sigma, St. Louis, MO) was dissolved in 100 mM sodium citrate buffer (pH 4.5). Mice were intraperitoneally injected with a single dose of 100 mg / kg. Three days after injection, fasting blood glucose was measured from the tail vein using a handheld blood glucose meter (One Touch UltraEasy; Lifescan). Then, they were deeply anesthetized with 140 mg / kg sodium pentobarbital and intracardially perfused with phosphate-buffered saline (PBS). Brains and livers were quickly taken out and immediately frozen in liquid nitrogen for further biochemical analyses.

**Culture and high glucose treatment of primary neurons and N2a and HEK293 cells**

To culture primary neurons, brains of embryonic day 18 (E18) Sprague-Dawley rats were quickly taken out and cortical neurons were [dissociated](javascript:;) and plated on matrigel-coated (BD Biosciences) glass coverslips, at approximately 100,000 cells / cm^2^, in Neurobasal medium (Invitrogen) containing 25 mM glucose with the supplement of B-27 (Invitrogen), 2 mM Glutamax-I (Invitrogen), and 2.5% fetal bovine serum (FBS, Hyclone). On the third day, cells were treated with the mitotic inhibitor 5-fluoro-2’-deoxyuridine (Sigma). On the 10^th^ day, the mature neurons were treated with 50 mM and 100 mM glucose. After 36 hours of treatment, the neurons were collected and homogenized using a polytron homogenizer in a radioimmunoprecipitation assay (RIPA) buffer (1% Triton X-100, 1% sodium deoxycholate, 0.2% sodium dodecyl sulfate (SDS), 0.15 M NaCl, 0.05 M Tris-HCl, pH 7.2) supplemented with complete mini protease inhibitor cocktail tablet (Roche, 11836170001) for protein detection using Western blotting.

N2a and HEK293 cells were cultured in Dulbecco’s modified Eagle medium (DMEM) containing 25 mM glucose with 10% FBS (Invitrogen) at 37°C with 5% CO_2_. After the cells reached 70-80% confluence in 6-cm dishes, they were treated with 50 mM (final in the medium, the same below) and 100 mM glucose (Sigma) for 6 hours. The osmotic pressure was adjusted with D-mannitol (Fisher). After the treatments, the cells were collected and homogenized using a polytron homogenizer in a RIPA buffer supplemented with complete mini protease inhibitor cocktail tablet (Roche, 11836170001) for protein detection using western blot analysis.

**Generation of conditional *Tpk* knockout mice**

To generate the conditional knockout of m*Tpk* allele (NM_013861.3), which has nine exons with the ATG start codon in exon 2 and TAA stop codon in exon 9, exon 4 was selected for targeting (Fig. S4A). The deletion of exon 4 results in the loss of function of *Tpk* gene by a frameshift. The detailed procedures are as follows.

Vector construction: mouse genomic fragments were amplified from the BAC clone with high fidelity Taq DNA polymerase and assembled into a targeting vector together with recombination sites and selection markers, as indicated on the vector map, in which the Neo cassette was flanked by Frt sites, cKO region flanked by LoxP sites, and DTA used for negative selection (Fig. S4B). The targeting construct was electroporated into C57BL/6 ES cells. Eighty-five (one 96-well plate) G418-resistant colonies were picked and screened by PCR using the strategy shown in Fig. S4C). The PCR primers were: 3’ arm - F: (GCTGACCGCTTCCTCGTGCTTTA) / 3’ arm - R: (GACCACCAAACAGCATACACCAGCT) and Loxp - F: (GTCACTTGAATATACCACAGTTCCCAG) / Loxp - R: (CATATTGGCACACTA CCCTGGAT). Two potential targeted clones (C10 and E7) were identified (Fig. S4D) and both were further analyzed by Southern blotting (Fig. S4E). The genomic DNA of the potential clones C10 and E7 were digested by NdeI and analyzed by Southern blotting for a 7.3-kb band from wild type allele and a 9.3-kb band from the recombinant allele using a probe generated by PCR with primers: 5’- probe, F_5’_: ATCTTTCCAGTGTCTCATTC / R_5’_: TACTTCTTGTTTCGGTCTT. They were also digested by XbaI and analyzed by Southern blotting for an 11.1-kb band from wild type allele and an 8.4-kb band from the recombinant allele, using a probe generated by PCR with primers: 3’- probe, F_3’_: GGATGTTGGCATTGAAGC / R_3’_：GATTTTAGGAGACGAGCA. Both analyses indicated that E7 was positive of recombination. The ES cells with the targeting confirmed by PCR and Southern analysis (E7) were injected into C57BL/6N blastocysts and subsequently transplanted into the uterus of pseudo-pregnant females. Chimeric males generated from albino-B6 blastocysts injected with the mutant ES cells were selected based on coat color and subsequently crossbred with C57BL/6N females. Germline mutant mice were crossbred with ROSA26::FLPe knock-in mice (Jackson Laboratory, stock no. 009086) to remove the FRT-flanked splice acceptor site, and neomycin resistance cassette. *Tpk^fl/+^* mice were identified using primers: LoxP_F: GAATTCCAAAGCCAGCATGT / LoxP_R: CCACTCACAGACCCTTTGCA and Neo F: ACTGTGCTTTGCTATCT CTACTGACA / Neo R: GGGCCACAAACACCTTCACATCT. The expected 192-bp fragment from wild type allele and 253-bp fragment from the recombinant allele represented positive F1 mice. Four positive pups (2, 4, 5, 6) from clone E7 were identified (Fig. S4F). The *Tpk^fl/+^* mice were interbred to generate homozygous mutant mice with the cKO potential (*Tpk^fl/fl^*). The *Tpk^fl/fl^* mice were crossed with excitatory neuron-specific Cre transgenic mice, *CaMKII-Cre^Ert2/+^* strain (EMMA ID: 02125) ^8^. To generate brain excitatory neuron-specific *Tpk*-cKO, the mice at 12 weeks of age were intraperitoneally injected with tamoxifen (50 mg/kg per day) for five consecutive days.

**Immunohistochemical staining**

After being deeply anesthetized with 0.14 g/kg sodium pentobarbital, the mice were intracardially perfused with PBS and then 4% paraformaldehyde for fixation. Brains were taken and embedded with OTC. Serial coronal sections (30 μm) were cut with a sliding microtome (Leica) and stained using a free-floating method. After being washed in PBS at pH 7.4, the sections were put into PBS containing 5% bovine serum albumin (BSA) and 0.5% Triton X-100 for 2 h at 37 °C in order to block non-specific reactions. Then, the sections were incubated with primary antibodies for NeuN (Millipore, MAB377, 1:500 or Abcom, ab104225, 1:500), GFAP (CST, 3670S, 1:500), Iba1 (Wako, 019-19741, 1:500 or Novus Biologicals, NB100-1028, 1:500), Anti-β-Amyloid 4G8 (Covance, SIG-39220, 1:50), p-Tau (Santa Cruz, sc-101815, 1:50), or TPK (Abcam, ab249546, 1:100) overnight at 4 °C. After being washed with PBS, the sections were incubated with goat anti-mouse antibodies conjugated to Alexa Fluor 488 (Invitrogen, 1:500) or Alexa Fluor 546 (1:500, Invitrogen) for 2 h at 37 °C. Nuclei were stained with DAPI (Sigma, D9542, 1:1000) and the slices mounted on 3-aminopropyltriethoxysilane (APES)-coated glass slides. Z-stack images were taken using a Nikon A1 (Tokyo, Japan) laser scanning microscope, with a 25x objective. Results were quantified using ImageProPlus (Media Cybernetics, Silver Spring, MD, USA). For detecting the effects of Aβ deposition on neuronal TPK expression, coronal brain slices were collected from 8-month-old APP/PS1 mice and immunostained with anti-TPK1, anti-NeuN antibodies. Amyloid plaques were labeled with thioflavin S. For each mouse, 50-80 field of views were acquired using a Nikon confocal microscope, with an oil immersion 63x objective (N.A. 1.4), on the somatosensory cortex region from 5-10 slices. In each field of view, neurons were identified and the distances between the neuronal cell body’s centroid point to the nearest plaque border were calculated. All neurons from each mouse were pooled together, and binned into different groups based on the distances to plaques. The number of neurons analyzed for each mouse were in a range of 1500-3000. Average measurements of TPK staining intensity were recorded from all the neurons in a group for each mouse. One-way ANOVA analysis was used to calculate the statistical comparison between different distance groups.

**Scanning electron microscopy**

Mice were intracardially perfused with PBS and then 2.5% glutaraldehyde for fixation. The brains were taken and the cortices of all mice were post fixed in 2.5% glutaraldehyde and stored in a refrigerator at 4 °C for 24 h. The tissues were cut into 2 × 2 × 5 mm cubes, washed three times with PBS for 30 min, and then soaked in 25% dimethylsulfoxide (DMSO) for 1 h and 50% DMSO for 12 h. Subsequently, the cortical tissues were freeze-cracked in liquid nitrogen, restored to 50% DMSO, and then washed four times with PBS as previously described ^9^. The samples were then washed with PBS three times for 30 min and post fixed with 1% osmium tetroxide, dehydrated with graded alcohol (25%, 30%, 50%,70%, 80%, 90%) once for each for 15 min, and substituted with 100% alcohol containing 1% isoamylacetate twice for 15 min each time. They were dried with liquid CO_2_ in an HCP-2 critical point dryer (Hitachi, Japan), and then coated with gold using Quorum Q150T ES (Quorum Technologies, United Kingdom) at 12 mA for 60 s. The metal-coated tissues were observed by a Gemin 300 field emission type scanning electron microscope (ZEISS, Germany) at 10 kV of accelerating voltage.

**Transmission electron microscopy**

Mouse cortical samples were trimmed into small cortical blocks and fixed in 4% paraformaldehyde and 2.5% glutaraldehyde in 0.1 M phosphate buffer (PB) for 48 hours. The fixed tissue blocks were washed twice in 0.1 M PB and then exposed to osmium tetroxide (1% in 0.1 M PB) for 30 min (protected from light). The samples were washed twice for 15 min in 0.1 M PB and three times in previously boiled ddH_2_O. Tissue blocks were then dehydrated for 15 min in 50% ethanol followed by exposure to 1% uranyl acetate in 70% ethanol for 40 min in the dark. The samples were further dehydrated in an ascending series of ethanol (95%, 100%, 100%) and propylene oxide before storage in Durcupan resin overnight at room temperature, followed by embedding in Durcupan resin and polymerization for 48 hours at 60 °C. The resin-embedded tissue blocks were cut into 70-nm-thick sections using a LEICA EM UC7 ultracut microtome equipped with a Jumbo Histo Diamond Knife (Diatome, Hatfield, PA, USA) and the sections collected onto copper formvar-coated grids. The grids were stained with lead citrate in a CO_2_-free environment for 2 min before imaging on a HITACHI H-7650 transmission electron microscope (TEM).

**Silver staining**

Brain slices were mounted on 3-aminopropyltriethoxysilane (APES)-coated glass slides and stained using *Bielschowsky* and Gallyas methods according to the manufacturers’ instructions (Bielschowsky Silver Stain Kit; ab245877; Abcam, USA; FD NeuroSilver™ Kit II; PK 301/301A; FD NeuroTechnologies, USA, respectively).

**Nissl staining**

Nissl staining of brain slices (7 µm thick) was performed according to the manufacturer’s instructions (Servicebio, G1036, China). Images were taken using Olympus VS120 Virtual Slide Microscope (VS120-S6-W, USA) with scan resolution at 10x (NA 0.4). The areas of brain slices were calculated using OlyVIA software.

**Protein purification and Western blotting of mouse brain tissues**

Brain tissues were lysed with the RIPA lysis buffer containing protease inhibitor mixture and PhosSTOP phosphatase inhibitor cocktail (Roche). Protein concentrations were determined with the Pierce™ BCA protein assay kit according to the manufacturer’s instruction (Thermo Scientific, Rockford, IL). A total of 10 mg of proteins were loaded in each lane on a 10% denaturing Tris/glycine gel for SDS-PAGE. For detecting CTFα and CTFβ, 50 mg of proteins were loaded in each lane on a 10% Tricine-SDS-PAGE. Proteins were transferred to polyvinylidene fluoride membranes (Millipore) and blocked in 5% milk in Tris-buffered saline supplemented with 0.1% Tween-20 (TBS-T, pH 7.4) for 2 h. The membranes were incubated overnight at 4 ^o^C with TBS-T containing 3% milk and primary antibodies for TPK (Abcam, ab249546, 1:1,000), Cleaved Caspase-3 (CST, 9664S, 1:1000), Cleaved PARP-1 (Beyotime, AG1043, 1:1000), p-NF-H (Biolegend, 801601, 1:1000), Tau (Tau46, CST, # 4019, 1:1,000), Phospho-Tau Ser396 (CST, # 9632S, 1:1,000), Phospho-Tau (Ser202, Thr205) Monoclonal Antibody (AT8, Invitrogen, MN1020, 1:1,000), Phospho-Tau Thr181(CST, #12885, 1:1,000), Phospho-Tau Thr205 (CST, #49561, 1:1,000), Total Tau (Abcam, ab80579, 1:1,000), APP (BOSTER, PB0101, 1:1,000), BACE1 (CST, 5606, 1:1000), Y188 (Abcam, ab220793, 1:1000), Insulin Receptor β (CST, # 3025, 1:1,000), IRS-1 (CST, # 3407, 1:1,000), Phospho-IRS-1 Ser612 (CST, # 3203, 1:1,000), Insulin Receptor β Tyr1150/1151 (CST, # 3024, 1:1,000), SLC19A3 (Proteintech,13407-1-AP, 1:1,000), SLC25A19 (Abclonal, A12373, 1:1,000), SLC19A2 (Bioss Antibodies, bs-10738R, 1:1,000), GAPDH (CST, # 2118, 1:1,000) and β-Actin (Proteintech, 60008-1-Ig, 1:10,000). Then, membranes were washed in TBS-T and incubated for 2 h with horseradish peroxidase (HRP)-conjugated anti-mouse or anti-rabbit IgG (Millipore) in TBS-T containing 3% milk. After a final wash in TBS-T, the membranes were incubated with enhanced chemiluminescence substrate (Pierce® Fast Western Blot Kit, 35050, USA) for 1–2 minutes, and signals were detected with Tanon 6200 Luminescent Imaging Workstation (Tanon Science & Technology Co., Shanghai, China). Results were quantified using Image J (N.I.H.).

**Real-time qPCR**

Total RNA was extracted from mouse brain tissues using the TRIzol reagent (Invitrogen). For Real-time RT-PCR experiments, complimentary DNA was synthesized using the PrimeScript™ RT reagent Kit with gDNA Eraser (TaKaRa, Japan) according to the manufacturer's protocol. Primers were designed using online Primer-BLAST software according to the instructions. Real-time PCR was performed using SYBR Green Master Mix (TaKaRa, Japan) on an ABI7500 Real-time PCR system (Applied Biosystems). All reactions were performed in triplicates, and the results were normalized to that of control groups from the same preparation.

| Primer | Forward | Reverse |
| --- | --- | --- |
| *β-Actin* | AGAAGGACTCCTATGTGGGTGA | CATGATCTGGGTCATCTTTTCA |
| *Il-1β* | GCAACTGTTCCTGAACTCAACT | ATCTTTTGGGGTCCGTCAACT |
| *Il-6* | GTCCTTCCTACCCCAATTTCCA | TAACGCACTAGGTTTGCCGA |
| *Tnf-α* | CCCTCACACTCAGATCATCTTCT | GCTACGACGTGGGCTACAG |
| *Gria*1 | CGAGTTCTGCTACAAATCCCG | TGTCCGTATGGCTTCATTGATG |
| *Gria*2 | AAAGAATACCCTGGAGCACAC | CCAAACAATCTCCTGCATTTCC |
| *Grin*1 | CGGCTCTTGGAAGATACAG | GAGTGAAGTGGTCGTTGG |
| *Grin*2b | TTTGGAGATGGGGAGATGG | CAGACACCCATGAAGCAATG |
| *mGluR*1 | AGTCTGCAGAACCGTCTGTG | GTTTACGGGACCTCTCAGGG |
| *mGluR*2 | GACTCTGGCTCCACTAAAGA | AGTCCTCACAAACACAGAGG |

**ELISA for Aβ42 and Aβ40**

The levels of Aβ42 and Aβ40 were determined by ELISA (Mouse Aβ42 or Aβ40 Colorimetric ELISA, KMB3441 or KMB3481, Invitrogen, Grand Island, NY, USA) according to the manufacturer’s instructions. In brief, brain tissues were weighed and homogenized using a polytron homogenizer in ice-cold PBS containing a protease inhibitor cocktail (Complete Protease Inhibitor Cocktail, Roche Diagnostics) followed by centrifugation at 16,000 x g for 20 min at 4°C. The supernatant was used for soluble Aβ determination. For insoluble Aβ, the pellets were further homogenized in a guanidine buffer (5 M guanidine HCl / 50 mM Tris-HCl, pH 8.0). The homogenates were mixed for 4 hours at the room temperature and then diluted 1:5 in PBS containing 5% BSA and 0.03% Tween-20 supplemented with the protease inhibitor cocktail followed by centrifugation at 16,000 x g for 20 min at 4°C. The supernatant was diluted and analyzed according to the manufacturer’s instructions. Final values of Aβ were expressed as pg per gram of brain tissue (wet weight).

**Behavioral tests**

Y-maze test

The Y-maze consists of 3 black horizontal arms (36 cm long, 5 cm wide, and 10 cm high) at 120° angles to each other. Mice were allowed to move freely through the Y-maze during a 5-min session. Alternation was defined as successive entries into the three arms on overlapping triplet sets. The percentage of alternations was calculated as the total number of alternations ×100 / (total number of arm entries - 2), which is not influenced by the unwillingness of the mouse to move. The performance was video-recorded and analyzed by an experimenter who was blinded to the genotypes of the animals using the image analyzing software (ANY-maze; Stoelting).

Nest-building behaviors

Mice were transferred to individual testing cages with one cotton rod. Two investigators who were blind to the grouping information independently assessed the scores of nesting (1–5 scores) according to the previous description ^10^ at 24 and 48 hours after entering the testing cages. The average values of the scores from two investigators were used as the final scores.

Morris water maze test

The Morris water maze test was performed as described previously ^11^. Briefly, the acquisition training paradigm for the Morris water maze consisted of eight trials (60 sec maximum; interval 30 min) each day for five consecutive days. Escape latency, path length, and velocity were recorded during training days. The probe test was performed 24 h after the last acquisition trial. The platform was removed, and mice were introduced into the water from a novel entry point. Mice were allowed to swim freely for 1 minute while the number of platform crossing, time spent in the target quadrant, and latency to the target quadrant were recorded.

Rotarod test

The rotarod treadmill (ENV-575, Med Associates, USA) was used to measure motor coordination and fatigue of the cKO and control mice. It consists of a computer-controlled stepper motor-driven drum with constant speed and accelerating speed modes of operation. The apparatus is divided into five test zones so that up to five animals could be tested at the same time. For accelerating speed test, the starting speed was 4 rpm and the speed accelerated from 4.0 to 40 rpm during 300 s. When the animal fell off the rotating drum, it broke a photobeam, leading to the automatic recording of the amount of time spent on the drum and the final speed.

Open field test

Open field arenas (MED-VOF-MS, Med Associates, USA) were used to assess anxiety phenotypes and general locomotor activities of the cKO and control mice. This system monitors locomotor activity using video cameras. Videos from the camera were displayed on the computer screen for user monitoring and recording for later review. Ambulatory and stereotypic behaviors, including ambulatory distance, time spent ambulatory, average velocity, and stereotypic time, were analyzed by the image analyzing software (ANY-maze; Stoelting).

**Magnetic resonance imaging (MRI)**

Mice were anesthetized with 1% isoflurane. MRI experiments were performed on a 9.4 T / 400 mm scanner (Agilent Technologies, Santa Clara, CA), using a quadrature conformal surface coil (7.5mm–12.5mm). Body temperature and respiratory rate were monitored and maintained throughout the experiment. A series of T2-weighted MR images were acquired and collected, typically along coronal orientation, using a RARE (Rapid Acquisition with Relaxation) of Field of view (FOV): 16 mm × 16 mm, matrix size = 192 × 192, slice thickness = 0.5 mm (28 slices, no gap), and bandwidth (BW) = 50 kHz. Acquisition parameters were as follows: repetition time (TR) = 5000 ms, echo spacing = 6.881 ms, Rare factor = 8, effective echo time (TE) = 27.52 ms, flip angle (FA) = 30°, number of averages (NA) = 5. A series of T1-weighted images were acquired using a FLASH (Fast Low Angle Shot) sequence with parameters of FOV: 16 mm × 16 mm, matrix size = 192 × 192, slice thickness = 0.5 mm (28 slices, no gap), BW = 60 kHz, TR = 350 ms, TE = 3 ms, FA = 30°, NA = 5. Brain volumes were analyzed using NIH ImageJ and MATLAB programs (MathWorks Inc., Natick, MA).

**Micro-PET scan:**

The radiolabeling synthesis of ^18^F-FDG was conducted as described previously ^12^. Before ^18^F-FDG injection, mice were fasted overnight with free access to water. Mice were injected with approximately 18.5 MBq (500 μCi) of ^18^F-FDG through the tail vein. Thirty minutes later, mice were anesthetized with isoflurane and PET/CT scanning was performed (Siemens Medical Solutions, Malvern, PA), which provided a 12.7 cm axial field of view and an intrinsic resolution < 1.7 ram. The entire static process included CT scanning for 10 min and PET scanning for 20 min. PET images were reconstructed using Fourier rebinning and 2-dimensional filtered back-projection (2D FBP) method (ramp filter and cutoff at Nyquist frequency) with an image matrix of 128 × 128 × 159, resulting in 3D images with a pixel size of 0.77 mm and a slice thickness of 0.796 mm. The images were analyzed using Inveon Research Workplace software (IRW, Siemens Medical Solutions, Malvern, PA). Regions of interest (ROIs) were automatically extracted from all micro-PET images using ^18^F-FDG murine brain templates of IRW. Relative FDG uptake was calculated for each ROI using the average tissue activity in the region.

**Intraperitoneal glucose tolerance test**

After fasting overnight, mice were injected intraperitoneally with glucose (2g / kg body weight). Blood glucose concentrations from tail vein were measured before glucose injection and 15, 30, 45, 60, 90 min after glucose injection using a handheld blood glucose meter (One Touch UltraEasy; Lifescan).

**Untargeted Metabolomics**

Mice were anesthetized with [pentobarbital](javascript:;) [sodium](javascript:;) and intracardially perfused with 4 °C PBS to remove blood. The brain was quickly dissected from the cranial cavity and immediately frozen in liquid nitrogen. The brain samples were stored at -80 °C and used within one month. To cortical samples of the cKO mice or control littermates (100 mg) were added 1 mL cold methanol / acetonitrile / H_2_O（2:2:1, v / v / v）and the mixture was adequately vortexed. The lysate was homogenized by an MP homogenizer (24 × 2, 6.0 M / S, 60 s, twice). The homogenate was sonicated on ice (twice, 30 min each time, with a 10-min interval) and then centrifuged for 20 min (14000 x g, 4 °C). The supernatant was collected and dried in a vacuum centrifuge. For LC-MS analysis, the samples were re-dissolved in 100 μL acetonitrile/water (1:1, v / v). Analyses were performed using a UHPLC (1290 Infinity LC, Agilent Technologies) coupled to a quadrupole time-of-flight (ABSciex Triple TOF 6600). For HILIC separation, samples were analyzed using a 2.1 mm × 100 mm ACQUIY UPLC BEH 1.7 µm column (Waters, Ireland). In both ESI positive and negative modes, the mobile phase contained A = 25 mM ammonium acetate and 25 mM ammonium hydroxide in water and B = acetonitrile. The gradient was 85% B for 1 min, linearly reduced to 65% B in 11 min, further reduced to 40% B in 0.1 min, and then kept for 4 min. After this, it was increased to 85% B in 0.1 min, with a 5 min re-equilibration period. For RPLC separation, a 2.1 mm × 100 mm ACQUIY UPLC HSS T3 1.8 µm column (Waters, Ireland) was used. In ESI positive mode, the mobile phase contained A = water with 0.1% formic acid and B = acetonitrile with 0.1% formic acid, while in ESI negative mode, the mobile phase contained A = 0.5 mM ammonium fluoride in water and B = acetonitrile. The gradient was 1% B for 1.5 min, linearly increased to 99% B in 11.5 min, and kept for 3.5 min. Then it was reduced to 1% B in 0.1 min and a 3.4 min of re-equilibration period was provided. The gradients were at a flow rate of 0.3 mL / min, and the column temperature was kept constant at 25 ℃. A 2 µL aliquot of each sample was injected. The ESI source conditions were set as follows: Ion Source Gas1 (Gas1) as 60, Ion Source Gas2 (Gas2) as 60, curtain gas (CUR) as 30, source temperature: 600 ℃, IonSpray Voltage Floating (ISVF) ± 5500 V. In MS only acquisition, the instrument was set to acquire over the m / z range 60-1000 Da, and the accumulation time for TOF MS scan was set at 0.20 s / spectra. In auto MS / MS acquisition, the instrument was set to acquire over the m / z range 25-1000 Da, and the accumulation time for product ion scan was set at 0.05 s / spectra. The product ion scan was acquired using information dependent acquisition (IDA) with high sensitivity mode selected. The parameters were set as follows: collision energy (CE) fixed at 35 V with ± 15 eV; declustering potentials (DP), 60 V (+) and −60 V (−); excluding isotopes within 4 Da; candidate ions to monitor per cycle at 10.

The raw MS data (wiff.scan files) were converted to MzXML files using ProteoWizard MSConvert before importing into freely available XCMS software. For peak picking, the following parameters were used: centWave m / z = 25 ppm, peakwidth = c (10, 60), prefilter = c (10, 100). For peak grouping, bw = 5, mzwid = 0.025, minfrac = 0.5. In the extracted ion features, only the variables having more than 50% of the nonzero measurement values in at least one group were kept. Compound identification of metabolites by MS / MS spectra was made with an in-house database established with available authentic standards. After normalization to total peak intensity, the processed data were uploaded before importing into SIMCA-P (version 14.1, Umetrics, Umea, Sweden), where they were subjected to multivariate data analysis, including Pareto-scaled principal component analysis (PCA) and orthogonal partial least-squares discriminant analysis (OPLS-DA). The 7-fold cross-validation and response permutation test was used to evaluate the robustness of the model. Metabolites of glycolysis and oxidative phosphorylation with p values less than 0.05 were considered as statistically significant.

**Measurement of TDP, thiamine monophosphate, and thiamine**

Fresh whole blood samples of mice were collected, anticoagulated with heparin, and deproteinized with 7.2% perchloric acid. The brain tissues were homogenized with 100 mM K_2_HPO_4_ (pH = 5.0) and deproteinized with isometric 6.4% perchloric acid. All samples were centrifuged and supernatants collected. The levels of TDP, thiamine monophosphate, and thiamine were measured as previously described ^11^.

**AAV vector production and intravenous injection.**

AAV.CAP-B10 vectors were generated according to the previous study ^13^ performed by BrainVTA (Wuhan, China), using the classical tri-transfection protocol. For AAV.CAP-B10.hsyn-TPK construct, mouse *Tpk* cDNA (NC_000072.7) was cloned into the AAV.CAP-B10 vector that contains a mature neuron specific promoter hSyn. Control and *Tpk*-targeting AAV.CAP-B10 viruses (AAV-TPK) expressed EGFP as an indicator. At the 5^th^ week after tamoxifen treatment, AAV-TPK and control vectors (200 µl total volume containing 2×10^11^ vector genomes) were injected to the *Tpk*-cKO mice and control littermates from the tail vein. Mice were used for behavior and pathological studies at the 4^th^ week after AAV injection (the 9^th^ week after tamoxifen treatment). TPK expression was validated by immunoblotting and immunohistochemical staining.

**Construction of N2a cell line with stable *Tpk* knockdown, *Tpk* overexpression, and treatment with TDP / thiamine**

The shRNA (ATCACTCCTGTGCCGATTATA) targeting *Tpk* was cloned into an integrated lentiviral vector. After transfection, N2a cells with the shRNA expression were selected and the RNAi mediated *Tpk* silencing was validated by Western blotting. The selected N2a cells were cultured in DMEM with 25 mM glucose supplemented with 10% FBS (Invitrogen) at 37 °C with 5% CO_2_. In order to examine the effects of *Tpk* overexpression and TDP / thiamine supplement on the levels of APP and BACE1, N2a cells, grown in 6-cm dishes to 70-80% confluency, were transfected with a plasmid encoding *Tpk* or treated with 0.1 mg / ml TDP (Sigma) / thiamine (Sigma). After 48 hours, the cells were collected and homogenized using a polytron homogenizer in a RIPA buffer with complete mini protease inhibitors (Roche, 11836170001) for detecting the levels of TPK, APP, and BACE1 proteins using western blot analysis.

**Statistical analysis**

Graphpad Prism 8 (version 8.01; GraphPad software) was used for statistical analyses. Student’s t-test for single comparisons or one-way ANOVA for multiple comparisons with appropriate Tukey’s or Dunnett’s Multiple Comparison tests or two-way ANOVA (for comparing two independent variables) followed by Bonferroni’s multiple comparisons test were used to determine statistical differences.

**Trial Design and Oversight**

Trial registration： ChiCTR1800014316 http://www.chictr.org.cn/ showproj.aspx?proj=24399; CTR20171631, http://www.chinadrugtrials.org.cn/.

We designed a 52-week, randomized, double-blind, placebo-controlled, multicenter phase II clinical trial to evaluate the efficacy and safety of benfotiamine plus donepezil in treating patients with mild-to-moderate Alzheimer’s disease, defined by the Mini Mental State Examination (MMSE) scores from 11 to 24 (inclusive; total scores ranging from 0 to 30, with lower scores indicating poorer cognition). The trial was conducted from Mar. 10, 2018, to Apr. 28, 2020, at 21 cognitive clinics of tertiary hospitals affiliated with Medical Colleges or Universities in China.

The protocol in accordance with Good Clinical Practice guidelines and the principles of Declaration of Helsinki was approved by the Committee of Medical Ethics of Huashan Hospital, Fudan University, the leading hospital of this trial, as well as by relevant local institutional review boards. All the participants lived in the community. Written informed consent was obtained from all participants and their legal caregivers prior to enrollment. The caregiver must be reliable and familiar with the patients, who spent a minimum of two hours per day and three days per week with the patient in order to observe the patents’ daily situations and accomplish the related scales.

The trial sponsor, Shanghai Raising Pharmaceutical Co., Ltd (Formerly known as Shanghai Rixin Biological Technology Co., Ltd.), designed and funded the trial, provided benfotiamine and placebo tablets, and purchased donepezil from Eisai company (Suzhou, China), analyzed the data, and aided in writing the manuscript. The first, third, and last authors wrote the first draft of the manuscript. All the authors vouch for the accuracy and completeness of the data, the fidelity of the trial to the protocol, and complete reporting of adverse events. All authors contributed to drafting and/or critical revision of the manuscript, reviewed and approved final versions of the manuscript for submission. The sponsor retained the right to review the manuscript for intellectual property purposes and to confirm the accuracy of all data and analyses. Confidentiality agreements are in place between the sponsor and the authors as well as between the sponsor and site investigators.

**Eligibility Criteria**

Participants were 50 to 80 years of age (inclusive), clinically diagnosed as Alzheimer’s disease according to the criteria of the National Institute on Aging and Alzheimer's Association (NIA-AA) in 2011 and had MMSE scores of 11 to 24 (inclusive), Clinical Dementia Rating (CDR) scores of 0.5 to 2.0 (inclusive; ranging from 0 to 3, with 0 indicating no dementia and 3 severe dementia), and atrophy of bilateral hippocampi represented by medial temporal lobe atrophy (MTA) scales of 2 to 4 (inclusive, ranging from 0 to 4, with 0 indicating no atrophy and 4 severe atrophy) identified in coronal brain MR images.

Participants must have been treated with a stable dose of donepezil (5 mg daily) for six months or longer before the enrollment and the same dose continued during the trial. The epidemiological data and medical history were obtained at the screening stage. Then, a battery of neuropsychiatric tests was performed, including MMSE, CDR, Hachinski Ischemic score (HIS, ranging from 0 to 18, with scores greater than 7 indicating a diagnosis of vascular dementia), and Hamilton Rating Scale for Depression-17 (HAMD-17, with scores over 17 indicating depression). The eligibility criteria included HIS scores of ≤ 4 and HAMD scores of ≤ 10 at the baseline. All the subjects were tested for blood hemoglobin, hepatic and renal functions, fasting blood glucose, glycosylated hemoglobin, creatine kinase, homocysteine, blood cholesterol and triglycerides, folate and vitamin B12 levels, antibodies of human immunodeficiency virus (HIV) and syphilis, as well as thyroid function, urinalysis, and electrocardiogram (ECG).

The exclusion criteria included: 1) The Fazekas Scale score is 3-point on MRI scanning (assaying the degree of brain white matter lesion, 0 to 3 scores, with a score of 3 representing severe lesion). 2) A diameter of cerebral infarction over 2 cm and any cerebral infarction in thalamus, hippocampus, entorhinal cortex, perirhinal cortex, angular cortex, and other grey matter nucleus found on MRI scanning. 3) Other brain diseases associated with cognitive impairment, e.g., dementia caused by cerebral vascular disease and other neurodegenerative diseases such as Lewy body dementia, frontotemporal dementia, and other clinical conditions including hydrocephalus, epilepsy, neuromyelitis optica spectrum disease, metabolic encephalopathy, toxic encephalopathy, various encephalitis, traumatic brain injury, and intracranial tumors. 4) Psychiatric disorders, such as schizophrenia, bipolar depression, delirium, and major depression. 5) Other conditions impacting cognitive abilities, including alcohol addiction, thyroid disease, hypoxia, folic acid and vitamin B12 deficiency. 6) Infectious diseases identified by positive reactions to HIV or syphilis antibodies in blood samples. 7) Significant abnormalities of laboratory tests: Blood alanine aminotransferase and creatine kinase up to twice the upper limit of normal (ULN), creatinine over 1.5 folds of ULN. 8) Poorly controlled hypertension or diabetes. 9) Heart rate less than 50 beats per minute; 10) Patients who had taken thiamine and its derivatives in recent 1 month; 11) Patients who had received memantine, acetylcholinesterase inhibitors except for donepezil (5 mg daily), or other nootropic drugs; 12) Blindness and deafness, which hindered subjects from completing all study requirements; 13) Severe conditions of gastrointestinal tract affecting drug absorption, including surgical procedures of gastrointestinal tract, dyspepsia, esophageal reflux, peptic ulcer, and gastric bleeding in recent six months. 14) Patients with other systematic disorders that were unsuitable to participate in this study as judged by their physicians.

**Randomization and masking**

After screening, participants entered a zero to 4-week introduction period in which they received monotherapy of donepezil (5 mg daily) before randomization in the study. Then, the participants were randomly assigned (1:1:1 allocation) to high-dose benfotiamine (600 mg daily) plus donepezil (5 mg daily, the same below, high-dose), low-dose benfotiamine (300 mg daily) plus donepezil (low-dose) or placebo plus donepezil (placebo) groups. To maintain a balanced distribution of case numbers and disease severity in the three groups, patients were stratified based on their disease severity assayed by CDR scores (mild, scores 0.5 and 1; moderate, score 2) and were randomized into the three groups according to the center-separate block randomization schedule.

The investigational drugs were oral placebo and / or benfotiamine tablets in a blister package delivered to the patients after randomization. The appearances of benfotiamine and placebo tablets were identical. Participants took the study drugs for 52 weeks and visited the investigative center once a month. All participants, their caregivers, the study sponsor, and study investigators at the centers were masked of the treatment assignment.

**Outcomes**

The primary outcome of efficacy was the change in the scores on the Alzheimer’s disease Assessment Scale-Cognitive Subscale 11 (ADAS-cog; ranging from 0 to 70, with higher scores indicating more severe cognitive impairment) from the baseline at the 52^nd^ week. All the scales were determined at the baseline (week 0) and repeated thereafter on weeks 12, 24, 36, and 52.

**Statistical analysis**

The changes of ADAS-cog scores at week 52 from baseline were used as the dependent variable of the primary endpoint. The ADAS-cog scores at baseline were adjusted using mixed methods for repeated measures (MMRM) model and the gate-keeping strategy was utilized for inter-group comparison. The differences between high-dose and placebo groups were first compared. If p value is less than 0.05 (inclusive) between high-dose and placebo groups (α = 0.05), then the differences between benfotiamine treatment (the combination of high-dose and low-dose groups) and placebo groups were compared. If that p value is also less than 0.05 (inclusive), the differences between low-dose and placebo groups were compared (α = 0.05). For more details, please see the Statistical Analysis Plan (SAP) in the attached supplementary materials.

The MMRM model included the fixed effects for treatment group, visit, the interaction of treatment group and visit, and the baseline value as a continuous covariate. The model used all available ADAS-cog scores data on weeks 12, 24, 36, and 52. An unstructured covariance structure was applied for MMRM analysis. In case the model did not converge with the unstructured covariance structure, the heterogeneous compound symmetry (CSH) would be used instead. Furthermore, if the model failed to converge with CSH, the heterogeneous Toeplitz structure (TOEPH) would be used. When the unstructured covariance structure was used, the denominator degrees of freedom were computed using the Kenward-Roger method. In case of other covariance structures, the BETWITHIN (SAS) option was used for the denominator degrees of freedom. Contrasts were constructed to compare each of the two benfotiamine treatment groups to placebo group at each visit. The least-squares (LS) mean change from baseline with the associated 95% CI was displayed for each treatment group. Estimated treatment differences (benfotiamine treatment vs. placebo) along with corresponding two-sided 95% CIs and p-values were also presented at each post-baseline visit. The missing data of the ADAS-cog scores were imputed using the last observation carried forward (LOCF).

**Post hoc analyses:**

Because the mean changes of ADAS-cog scores at week 52 from baseline exhibited a tendency of dose-dependent effect but no statistical difference between high-dose, low-dose, and placebo groups, we further performed an ad-hoc analyses to examine the changes of ADAS-cog scores at week 52 from baseline in placebo group based on the disease severity ranked by the MMSE scores at baseline. The results showed that in placebo group, the changes of ADAS-cog scores were not significant in patients with the MMSE scores of 20 to 24 but exhibited obvious increases in patients with the MMSE scores of 11 to 19 (Extended Data Fig. 15). Therefore, we further divided the participants into two subgroups: mild subgroup with the MMSE scores of 20 to 24 (inclusive) and moderate subgroup with the MMSE scores of 11 to 19 (inclusive) at baseline.

Analyses for the changes of ADAS-cog scores in the subgroups of moderate cases were first performed to examine the differences between high-dose and placebo groups using the same MMRM model described as stated above. This model will include the same fixed effects.
